# Biological Insights from Schizophrenia-associated Loci in Ancestral Populations

**DOI:** 10.1101/2024.08.27.24312631

**Authors:** Tim B. Bigdeli, Chris Chatzinakos, Jaroslav Bendl, Peter B. Barr, Sanan Venkatesh, Bryan R. Gorman, Tereza Clarence, Giulio Genovese, Conrad O. Iyegbe, Roseann E. Peterson, Sergios-Orestis Kolokotronis, David Burstein, Jacquelyn L. Meyers, Yuli Li, Nallakkandi Rajeevan, Frederick Sayward, Kei-Hoi Cheung, Project Among African-Americans to Explore Risks for Schizophrenia (PAARTNERS), Consortium on the Genomics of Schizophrenia (COGS), Genomic Psychiatry Cohort (GPC) Investigators, Lynn E. DeLisi, Thomas R. Kosten, Hongyu Zhao, Eric Achtyes, Peter Buckley, Dolores Malaspina, Douglas Lehrer, Mark H. Rapaport, David L. Braff, Michele T. Pato, Ayman H. Fanous, Carlos N. Pato, PsychAD Consortium, Cooperative Studies Program (CSP) #572, Million Veteran Program (MVP), Grant D. Huang, Sumitra Muralidhar, J. Michael Gaziano, Saiju Pyarajan, Kiran Girdhar, Donghoon Lee, Gabriel E. Hoffman, Mihaela Aslan, John F. Fullard, Georgios Voloudakis, Philip D. Harvey, Panos Roussos

## Abstract

Large-scale genome-wide association studies of schizophrenia have uncovered hundreds of associated loci but with extremely limited representation of African diaspora populations. We surveyed electronic health records of 200,000 individuals of African ancestry in the Million Veteran and All of Us Research Programs, and, coupled with genotype-level data from four case-control studies, realized a combined sample size of 13,012 affected and 54,266 unaffected persons. Three genome-wide significant signals — near *PLXNA4*, *PMAIP1*, and *TRPA1* — are the first to be independently identified in populations of predominantly African ancestry. Joint analyses of African, European, and East Asian ancestries across 86,981 cases and 303,771 controls, yielded 376 distinct autosomal loci, which were refined to 708 putatively causal variants via multi-ancestry fine-mapping. Utilizing single-cell functional genomic data from human brain tissue and two complementary approaches, transcriptome-wide association studies and enhancer-promoter contact mapping, we identified a consensus set of 94 genes across ancestries and pinpointed the specific cell types in which they act. We identified reproducible associations of schizophrenia polygenic risk scores with schizophrenia diagnoses and a range of other mental and physical health problems. Our study addresses a longstanding gap in the generalizability of research findings for schizophrenia across ancestral populations, underlining shared biological underpinnings of schizophrenia across global populations in the presence of broadly divergent risk allele frequencies.

## Introduction

Schizophrenia and related psychoses occur in all human populations, but are diagnosed most frequently among those racialized as Black and those of primarily African ancestries^1^. In the United States, such patients experience worse treatment outcomes (e.g. hospitalization, functioning) as a consequence of racism and other social determinants of health, compounding pervasive inequities which impact morbidity, mortality, and quality of life^2,3^. Decades of research point resoundingly to a heritable basis of risk, which is shared across diagnoses and populations^4,5^ and does not account for differences in prevalence and presentation^6,7^. Furthermore, because the largest genome-wide association studies (GWAS) have overwhelming recruitment bias towards European populations^8^, the continued underrepresentation of African ancestries in genomics research threatens to limit the benefits of novel biological insights and advances in precision medicine, further entrenching these disparities.

A primary obstacle in genomics research on schizophrenia is its extreme polygenic architecture. Hundreds of loci have been identified to date, which exert very small effects in concert with thousands more that do not attain stringent genome-wide significance. Importantly, prior research indicates that most of the currently attributable risk (i.e. in European populations) resides in variation predating human migrations out of Africa, and is shared across worldwide populations. However, diverging allele frequencies, effect sizes, and patterns of linkage disequilibrium (LD) render polygenic risk scores (PRS) that decline continuously in performance as genetic distance between training and target samples increases^9–11^. These limitations are especially marked in populations of African ancestry.

Building on our recent work characterizing and validating psychiatric diagnoses from electronic health records (EHRs)^12,13^, we newly integrate genome-wide single nucleotide polymorphism (SNP) genotyping data for over 124,000 African ancestry individuals (AFR) from the Million Veteran Program (MVP)^14^ and Cooperative Studies Program (CSP) #572^15^. We comprehensively assess the replicability of major schizophrenia findings and, for the first time, recapitulate characterizations of the genetic architecture that to date have been limited to populations of European (EUR) and East Asian (EAS) ancestry. Combining results with civilian data from the All of Us (AOU) Research Program^16^, and four published case-control studies^17–20^ culminated in a multi-stage GWAS of 13,012 affected persons and 54,266 controls of admixed African ancestries (**Supplementary Note**). Subsequent meta-analysis with European and East Asian data yielded a total of 86,981 cases and 303,771 controls. An overview of the AFR-specific and cross-ancestry meta-analysis strategies is given in **Figure 1A**. To increase our understanding of the biological mechanisms underlying schizophrenia across ancestries, we integrated outcomes from the cross-ancestry fine-mapping analysis with single-cell functional genomic data from the human dorsolateral prefrontal cortex.

**Figure 1.**
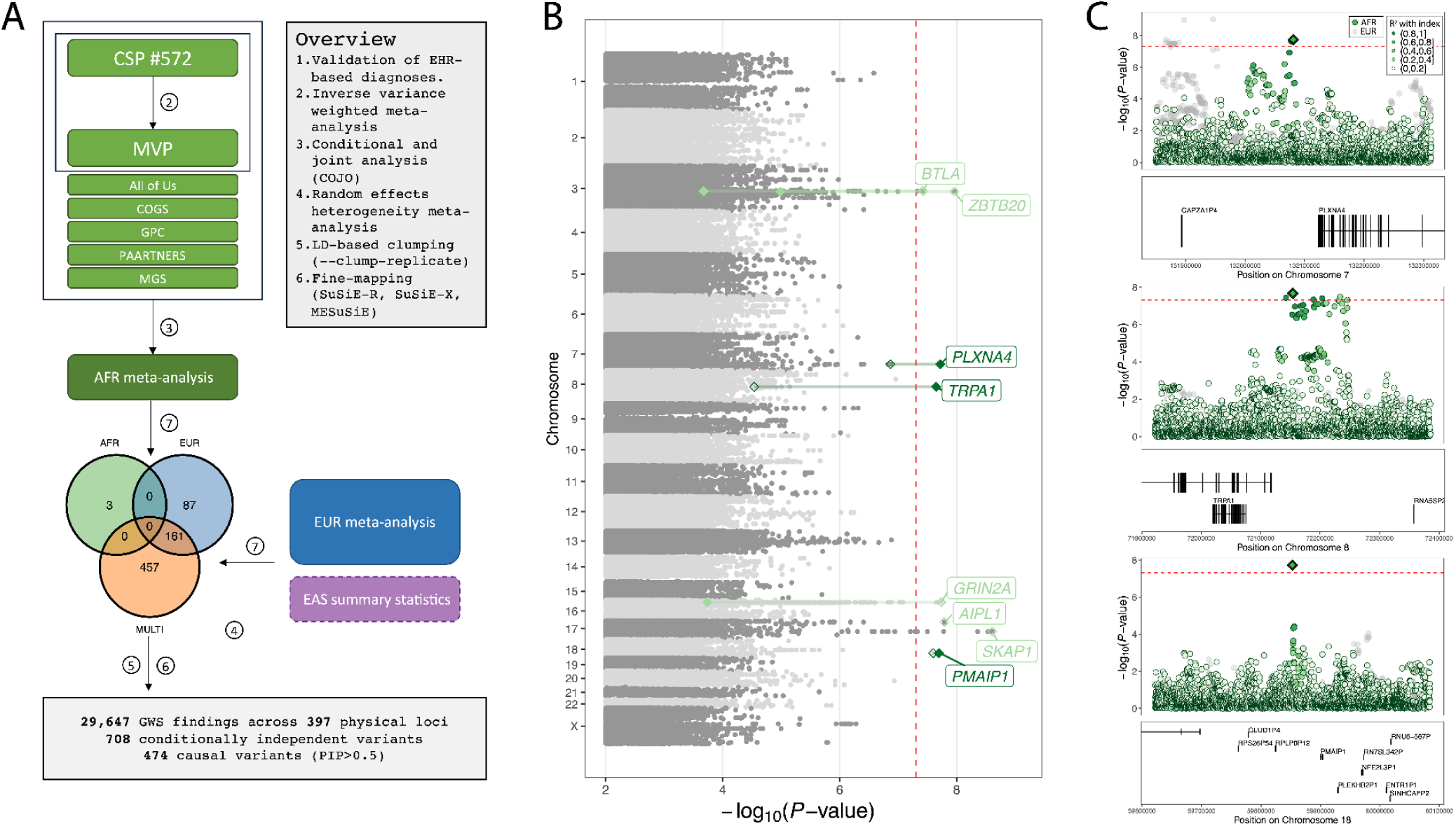
Overview of AFR and cross-ancestry meta-analyses. **(A)** Stages 1 and 2 of AFR meta-analysis, and incorporation of summary statistics for EUR and EAS, with summaries of GWAS variants, conditionally independent signals, causal variants, and physical loci. **(B)** Manhattan plot of discovery (Stage 1) AFR GWAS of schizophrenia in CSP #572 and MVP. Loci achieving genome-wide significance in discovery (Stage 1) (light green text) and meta-analysis (Stage 2) (dark green text) stages are highlighted; empty and filled diamonds represent the corresponding P-values in each stage. Index variants absent in AFR meta-analysis are displayed by an empty diamond. **(C)** Regional association plots for the three genome-wide significant loci in the AFR meta-analysis; (green dots, where the shade represents the strength of LD with the index variant (a diamond); corresponding results for EUR meta-analysis are displayed (grey dots).

## Results

### Schizophrenia genetic associations in AFR populations

We initially compared 6,408 patients with schizophrenia and 20,065 controls enrolled in CSP #572 and MVP (Stage 1), and then extended our analysis to an additional 6,604 cases and 34,201 controls (Stage 2) from AOU^16^, Consortium on the Genetics of Schizophrenia (COGS)^18^, Genomic Psychiatry Cohort (GPC)^20,21^, Molecular Genetics of Schizophrenia (MGS)^19^, and Project among African-Americans to explore risks for schizophrenia (PAARTNERS) studies^17,22^ (**Extended Data Table 1; Supplementary Data 1**).

Our GWAS in CSP #572 and MVP (Stage 1) uncovered six statistically independent genome-wide significant (*P*<5×10^-8^) associations with SNPs in *ZBTB20* on 3q13.31, *BTLA* on 3q13.2, *GRIN2A* on 16p13.2, near *WSCD1* and *AIPL1* on 17p13.2, downstream of the Homeobox B (*HOXB*) gene cluster on 17q21.32, and upstream of *PMAIP1* on 18q21.32 (**Figure 1B**; **Extended Data Table 2**). Of these, only *PMAIP1* was directly replicated in the expanded AFR meta-analysis (13,012 cases and 54,266 controls), while two additional regions — upstream of *PLXNA4* on 7q32.3 and *TRPA1* on 8q21.11 — attained genome-wide significance with this analysis (**Figure 1B**; **Figure 1C**).

Notably, none of the specific genome-wide significant associations observed in AFR have been reported in published studies of EUR or EAS populations. However, suggestive AFR findings (*P*<10^-5^) included previously replicated associations with SNPs in *FAM120A* (9q22.31) and *DCC* (18q21.2) (**Supplementary Table 1**).

### Cross-ancestry GWAS of schizophrenia

We obtained EUR summary statistics from the Psychiatric Genomics Consortium (PGC3; 53,386 cases and 77,258 controls) and combined these with new MVP-EUR results (6,579 cases and 155,490 controls) to yield an expanded GWAS of 59,965 cases and 232,748 controls (**Figure 2A - left panel**; **Extended Data Table 1**). We then combined our AFR results (13,012 cases and 54,266 controls) with these expanded EUR results and available EAS summary statistics^23^ (14,004 cases, 16,757 controls) in cross-ancestry meta-analyses across 18,982,014 individual variants in 86,981 cases and 303,771 controls (**Figure 2A - right panel; Extended Data Table 1; Supplementary Data 1**).

**Figure 2.**
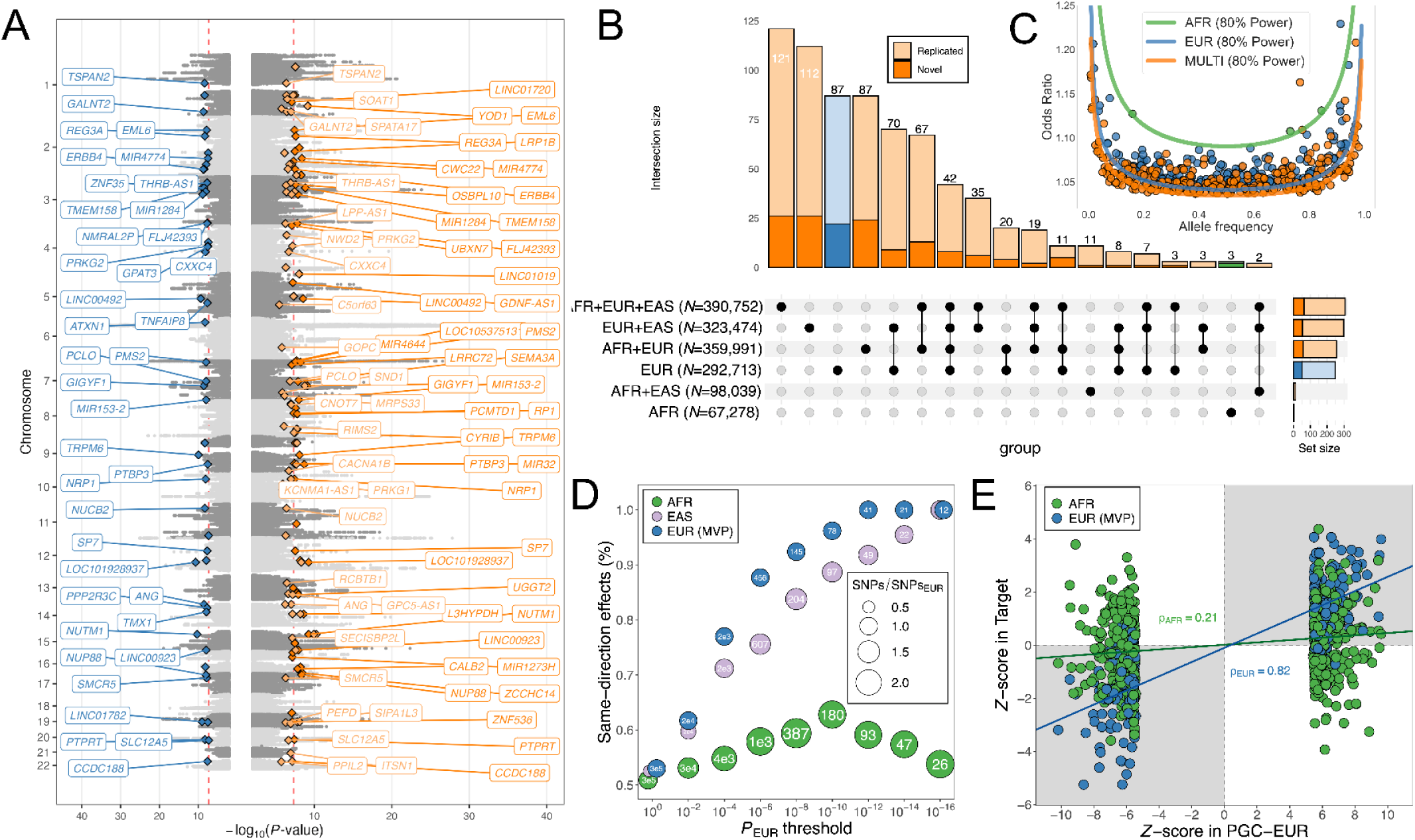
Cross-ancestry schizophrenia meta-analyses. **(A)** Miami plot displaying expanded EUR results (*left*; 59,965 cases and 232,748 controls) and cross-ancestry findings (*right*; 86,981 cases and 303,771 controls). Each conditionally independent lead variant within 1Mb is displayed as a diamond; novel findings in expanded EUR meta-analyses (blue); novel findings in cross-ancestry GWAS (orange); novel variants in analyses of two ancestries are also shown (lighter shades). **(B)** Upset Plot displaying the distribution of distinct index SNPs across updated EUR meta-analysis and cross-ancestry meta-analyses. The total number of conditionally independent SNPs for each are displayed in the lower left panel. Single-ancestry (blue) EUR versus cross-ancestry (orange) meta-analyses are highlighted. **(C)** Frequencies and Odds Ratios for COJO SNPs in EUR, AFR, and cross-ancestry meta-analyses, with respect to the alternative (tested) allele. Corresponding 80% power lines are displayed. **(D)** Comparison of directional concordance across single-ancestry meta-analysis results. **(E)** Standardized effect estimates for MVP-EUR and AFR at genome-wide SNPs in PGC3-EUR, with regression lines; panels shaded in grey contain findings with directionally concordant effects.

We resolved 19,529 genome-wide significant associations (excluding 5,021 mapping to the extended MHC region) to 307 statistically independent signals using conditional and joint (COJO) multi-SNP analysis (**Methods**), which accounts for linkage disequilibrium (LD) among nearby variants (**Supplementary Table 2**). By comparison, meta-analyses of EUR and EAS, EUR and AFR, and AFR and EAS yielded 298, 257, and 13 independent signals, respectively (**Supplementary Tables 3-5**). Taken together with within-ancestry results for AFR (3) and EUR (248) (**Supplementary Table 6**), this corresponded to 708 unique variants spanning 397 physically distinct chromosomal loci (**Figure 2B**). As expected, there was starkly lower power to detect genome-wide significant associations in AFR compared to EUR populations and only marginally greater power in cross-population analyses of three ancestries (**Figure 2C**).

We intersected our findings with 270 loci in the most recent PGC study^24^ using LD-”clumped” intervals, highlighting 151 independent variants in 111 novel loci that were more than 1Mb from a previously reported locus (**Figure 2A** and **Figure 2B**; **Supplementary Table 7**). Multi-ancestry GWAS findings revealed additional novel loci implicating genes, including *CACNA1B*, *KCNN4-AS1*, *NRP1*, *PTPRT*, *SEMA3A*, and *SLC12A5*, which play important functions in neuronal signaling.

We compared directions of allelic effects between ancestries, observing that, for progressively more stringent *P*-value thresholds (*P*_T_) applied to the PGC-EUR findings, the proportion of same-direction effects in EUR replication and EAS results converged to 1 (**Figure 2D**). As applied to AFR results, in which the same PGC-EUR signals corresponded to a larger effective number of LD-independent SNPs, we observed fractions of same-direction tests exceeded 60% at (*P*_sign_<10^-3^ at *P*_T_<10^−10^); however, this fraction exceeded the null expectation of 50% concordance at all tested thresholds. A direct comparison of LD-independent genome-wide significant SNPs revealed a modest but significant correlation between AFR and EUR effects (ρ=0.209; *P*=7.55×10^-7^), despite fewer directionally concordant results than seen with an independent EUR sample (ρ=0.812; *P*=8.25×10^−53^) (**Figure 2E; Supplementary Table 8**).

### Enhanced fine-mapping of schizophrenia-associated loci

We evaluated the impact of expanded diversity on fine-mapping resolution at new and previously associated loci. From the COJO SNPs in each meta-analysis, we defined 773 unique intervals separated by at least 250 kilobases (kb), represented by the most significant index SNP in a given window (**Supplementary Table 9; Supplementary Data 2-4**). For each window, we estimated a 95% credible set of SNPs, corresponding to a genomic interval containing one or more causal variants driving a given association signal. For the EUR-only and the meta-analyzed GWAS, we used SuSiE-R with a “blended” LD matrix (**Figure 3A**; **Methods**). The number of causal SNPs we identified (posterior inclusion probability, PIP>0.5 and *P*<5×10^-8^) was 51 for EUR only, 245 for AFR+EUR, 83 for EUR+EAS, and 366 for AFR+EUR+EAS (**Supplementary Table 10**). Additionally, fine-mapping between the unique pairs of distinct ancestries (AFR+EUR, EUR+EAS, AFR+EAS) was performed using MESuSiE (**Figure 3A**; **Supplementary Tables 11-18**; **Methods**). Lastly, for the three distinct ancestries (AFR+EUR+EAS), fine-mapping was conducted with SuSiEx (**Figure 3A**; **Methods**), which is less conservative than MESuSiE as it considers all comparisons among ancestries. The number of SNPs with PIP > 0.5 was 248 for AFR+EUR, 41 for EUR+EAS, 68 for AFR+EAS, and 261 for AFR+EUR+EAS (**Supplementary Tables 19-20**).

**Figure 3.**
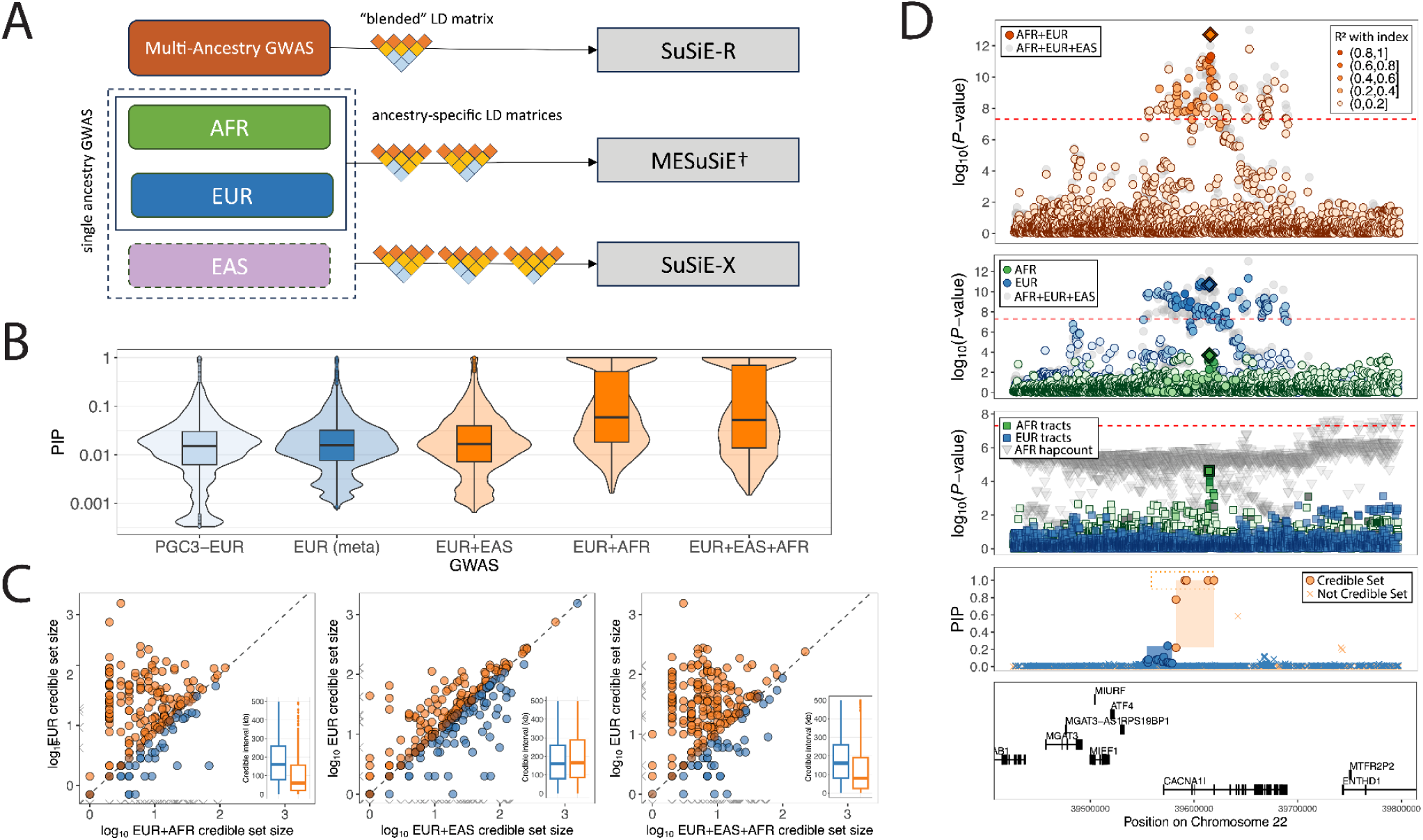
Improved fine-mapping resolution in cros-ancestry schizophrenia GWAS. **(A)** Overview of fine-mapping approaches employed and their particular handling of cross-ancestry LD information. **(B)** Distribution of PIP values for SNPs in credible sets identified for PGC-EUR (6,234), EUR meta-analysis (8,066), EUR and EAS (7,931), EUR and AFR (2,514), and cross-ancestry meta-analysis of EUR, EAS, and AFR (3,467). Values are transformed to the log_10_-scale **(C)** Comparative improvements in fine-mapping resolution from cross-ancestry versus EUR-only analyses. Each point represents a credible set of SNPs in a given meta-analysis; points above the dotted line indicate a smaller credible set size in the trans-ancestry meta-analysis; grey points along axes indicate signals which SuSiE-R did not detect as causal. Inset panels display these distributions as split violin plots. (**D**) Regional findings at *CACNA1I* with EUR with EUR+AFR meta-analysis results (*upper*), and deconvolved AFR tracts and haplotype counts (*lower*).

We observed higher posterior probabilities following meta-analysis of EUR with EAS and AFR results (**Figure 3B**) and shorter genomic intervals (kb spanned by credible SNPs) (**Figure 3C**). In the EUR only analysis, out of 773 unique intervals separated by at least 250 kb, 606 indexes had at least one credible set defined, with 72 SNPs exhibiting a PIP greater than 0.5 and 51 SNPs reaching genome-wide significance (*P*<5×10^-8^). In the AFR+EUR analysis, 671 unique indexes were identified, including 1,056 SNPs with a PIP greater than 0.5 and 245 SNPs that were genome-wide significant. For the EUR+EAS analysis, we found 678 unique indexes with at least one credible set, among which 208 SNPs had a PIP greater than 0.5 and 83 SNPs were genome-wide significant. Lastly, the AFR+EUR+EAS analysis revealed 697 unique indexes, with 1,353 SNPs having a PIP greater than 0.5 and 366 SNPs reaching genome-wide significance.

### Leveraging local ancestry in admixed AFR populations

We investigated the impact of admixture in AFR individuals through local ancestry deconvolution, followed by joint modeling of AFR and EUR haplotype tracts using an adaptation of the *tractor* approach^25^ (**Methods**). We observed extended genomic regions that harbored significant (*P*<10^-6^) associations between the number of AFR haplotypes carried by an individual and reduced disease risk (**Extended Data Figure 1A**). Comparing standardized effect sizes estimated from AFR tracts and EUR tracts to PGC-EUR results (*P*_T_=5×10^-8^), we observed a stronger correlation with EUR tracts (ρ=0.435; *P*=1.88×10^−11^) than AFR tracts (ρ=0.263; *P*=4.29×10^−10^) (**Extended Data Figure 1B**). Similarly, the proportion of same-direction tests was greater for EUR (0.68; *P*_sign_=8.67×10^-8^) than AFR (0.58; *P*_sign_=2.35×10^-3^) tracts (**Extended Data Figure 1C; Supplementary Table 8**).

Examination of tract-level results revealed distinct AFR-specific evidence of association near *PLXNA4* (*P*_AFR_=6.75×10^-5^; *P*_EUR_=0.999), *PMAIP1* (*P*_AFR_=7.22×10^-5^; *P*_EUR_=0.985), and *TRPA1* (P_AFR_=5.56×10^-4^; *P*_EUR_=0.207) (**Extended Data Figure 2A-C**). We observed even stronger tract-level associations with SNPs in *CLEC16A* on 16p13.13 (*P*_AFR_=3.94×10^-7^; *P*_EUR_=0.6414), *DOCK4* on 7q31.1 (*P*_EUR_=1.15 ×10^-6^; *P*_AFR_=0.649), and *GRIN2B* on 12p13.1 (*P*_AFR_=8.93×10^-8^; *P*_EUR_=0.696). At *GRIN2A*, we observed overlapping but distinct associations based on AFR (*P*_AFR_=6.10×10^-6^; *P*_EUR_=2.35×10^-3^) and EUR tracts (*P*_EUR_=1.42×10^-6^; *P*_EUR_=0.884) (**Extended Data Figure 2D-F**).

Taken together, these results suggest the intriguing possibility of population-specific haplotypes or complex patterns of LD that hinder detection of causal schizophrenia-related variation in the presence of local admixture. We explored the potential for enhanced fine-mapping resolution (i.e. beyond standard trans-ancestry meta-analysis) by re-estimating credible sets based on meta-analysis of our overall EUR findings with tract-level AFR statistics. In particular, at the *CACNA1I* locus, this approach narrowed the corresponding credible set from 64.7kb to 39kb spanning exons 2 and 3 and introns 1-3 (**Figure 3D)**. Within this region, 10 SNPs had a PIP greater than 0.9, compared to a maximum PIP of 0.244 based on EUR results alone.

### Comparative estimates of SNP-based heritability

We sought to better characterize the population-specific genetic architectures of schizophrenia in AFR and EUR populations, by leveraging available individual-level genotype data for the combined CSP #572 and MVP cohorts. Using common SNPs with MAF ≥ 1%, we obtained near-identical estimates of SNP-*h*^2^ for AFR (0.183±0.013) and EUR (0.183±0.017) participants (**Extended Figure 3A; Supplementary Tables 21-22**); including variants with MAF greater than 0.1% yielded comparable increases in SNP-*h*^2^ for AFR (0.268±0.026) and EUR (0.270±0.019). Adjusting these estimates by the number of SNPs in each population illustrates that, in EUR populations, an equivalent fraction of variability in individual risk is explained by fewer SNPs (**Extended Data Figure 3A**). In contrast, calculating SNP-*h*^2^ from ∼1.2M HapMap3 variants yielded markedly attenuated estimates for AFR (0.138±0.014) but not EUR (0.196±0.012), highlighting the limitation of using “convenience sets” of SNPs optimized for EUR populations ^26–28^.

Given the extensive genetic diversity present in AFR populations, we partitioned SNPs which were common (MAF>0.1%) in AFR individuals but rare (MAF<0.1%) in EUR, and variants which were common in both. Among AFR individuals, shared common genome-wide variants explained around a quarter of the total attributable variance (0.074±0.015), while less common (MAF<1%) variation explained a 1.5-fold larger fraction of individual variability in liability (0.114±0.027) (**Extended Figure 3B; Supplementary Tables 23**).

Next, we partitioned SNPs in EUR participants based on the absolute allele frequency difference (AFD) ^29^ with AFR populations. We found that a negligible proportion of variance was explained by population-”specific” variation (0.014±0.029), with most of the attributable variance explained by SNPs in the third (0.075±0.016) and fourth (0.071±0.013) AFD quartiles (**Extended Figures 3C; Supplementary Table 24**). Taken together with the observations above, this suggests that variation exhibiting greater divergence between populations harbors a larger fraction of disease-related variance.

### Cell-type convergence of EUR and AFR heritability for schizophrenia

Previous studies support the enrichment of EUR schizophrenia risk loci for gene expression markers related to excitatory and inhibitory neurons^24,30,31^. We examined whether the genetic architecture of schizophrenia in AFR converges to similar cell type enrichment. For this purpose, we used the extensive PsychAD single-cell atlas of 6.3 million nuclei from the human prefrontal cortex^32^ and applied the scDRS method^33^ that considers transcriptional heterogeneity between individual cells to calculate polygenic risk score for each cell population (**Methods**). Consistent with multiple lines of prior evidence ^24,30,31^, we observed a significant enrichment in all subtypes of excitatory and inhibitory neurons in EUR (**Figure 4B**). While the EUR and AFR enrichments are well correlated (Pearson’s *r* = 0.702; **Figure 4C**), not all significantly enriched neuronal subtypes were replicated in AFR which can be attributed to lower GWAS power in AFR. Out of the significant findings, oligodendrocyte precursor cells (OPCs) were more highly enriched in AFR than EUR GWAS (**Figure 4B**).

**Figure 4.**
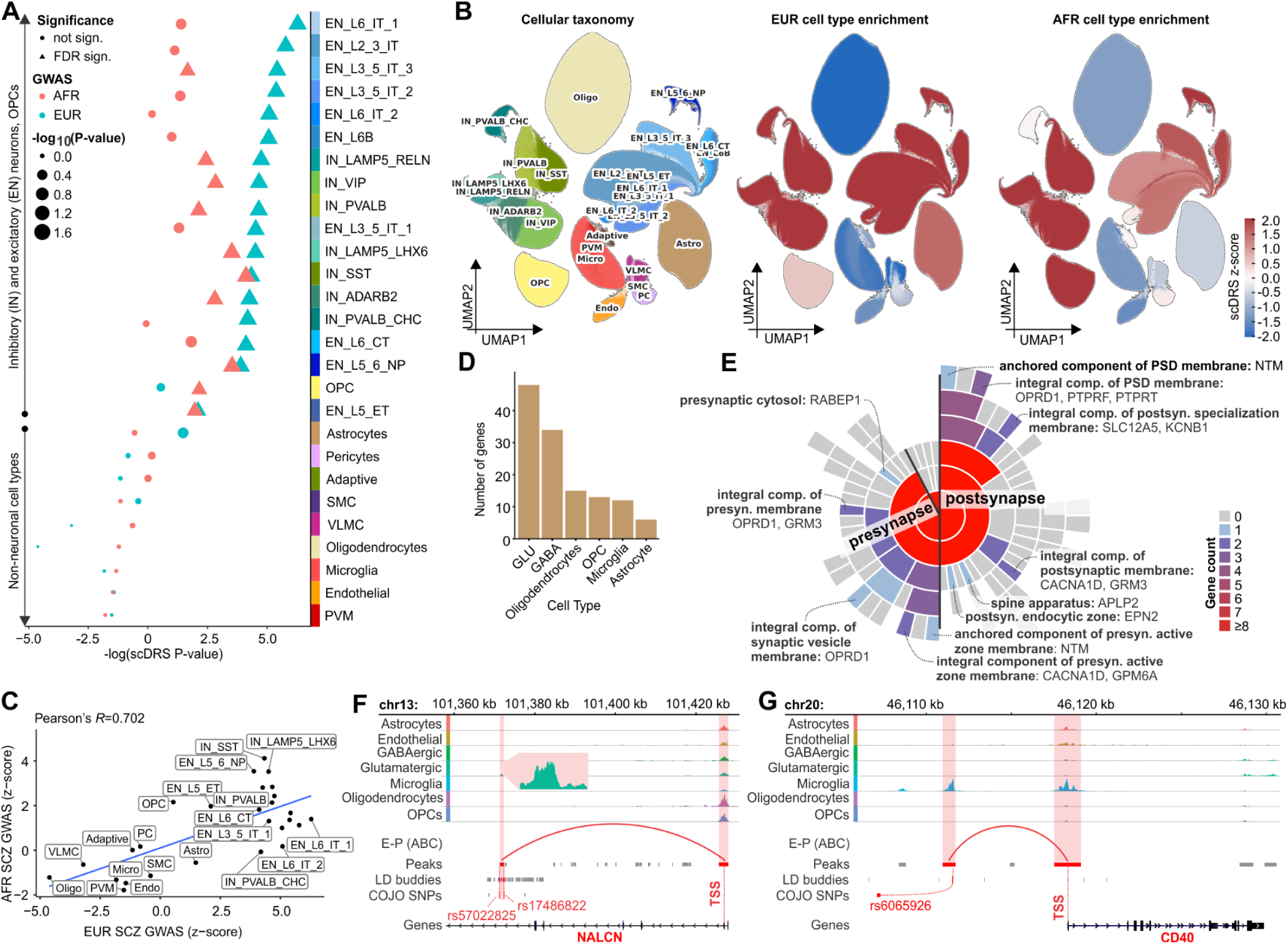
Cell-type specific heritability patterns and disease regulation in schizophrenia. **(A)** Enrichment of EUR and AFR for 27 cell subtypes from the single cell atlas of the prefrontal cortex. **(B)** UMAP visualizations of single cell atlas colored by the cell taxonomy (left), EUR enrichment (middle) and AFR enrichment (right). IN: inhibitory neurons / GABAergic, EN: excitatory neurons / glutamatergic, SMC: smooth muscle cells, VLMC: vascular leptomeningeal cells, PVM: Perivascular macrophages. **(C)** Correlation of scDRS z-score across cell subtypes between EUR and AFR. **(D)** Number of causal genes per cell type, stratified by E-P_ABC-MAX_, TWAS, and shared nominations (top). Shared nominations are also displayed separately in the bottom plot for better clarity. **(E)** Mapping of prioritized genes in the synaptic structure using the SynGO database^46^. The sunburst plot positions the synapse at its core, with layers for pre- and post-synaptic regions in the first ring, followed by specific categories in outer rings. The color coding represents the gene count for each category. **(F-G)** Normalized snATAC-seq pseudobulk tracks demonstrating the cell-specific regulation of the NALCN (E) and CD40 (F) affected by GWAS COJO SNPs rs6065926 and rs17486822/rs57022825

### Cell-type transcriptome convergence of EUR and AFR heritability for schizophrenia

We leveraged single-nucleus gene expression data from the prefrontal cortex of 920 EUR and 321 AFR donors that are included in the PsychAD cohort^32^ to perform an ancestry-stratified transcriptome-wide association study (TWAS). Across 31 cell-type-specific models, EUR models could confidently impute 18,219 unique genes across 112,474 gene-cell-type combinations, while AFR models could confidently impute 16,956 unique genes across 74,002 gene-cell-type combinations (**Methods**). Integrative analysis of the 31 cell-type-specific EUR models with the expanded EUR GWAS identified 2,057 unique genes (Bonferroni-adjusted *P*-value < 0.05). Similar analysis using the AFR models with the AFR GWAS identified only 3 unique genes at the same statistical threshold, likely due to the lower AFR GWAS power. To assess concordance between EUR and AFR schizophrenia TWAS, we quantified the gene correlation as a function of top ranked genes (as defined by meta-analysis *P*-value). Across all well-represented cell-types, we find that correlation among EUR and AFR TWAS is ∼0.7-0.9 for the top ranked genes (**Extended Data Figure 4**), highlighting the biological convergence capturing genetically driven schizophrenia-associated gene expression shared between EUR and AFR.

### Identification of putative causal genes through enhancer-promoter interaction model

Expression quantitative trait loci (eQTL) mapping and colocalization with GWAS variants is a common method for nominating credible causal genes^34^; however, eQTLs explain only a small fraction of GWAS signals due to their clustering near transcription start sites and association with genes that have simpler regulatory landscapes^35^. To address this limitation, cell-specific enhancer-promoter (E-P) models^36^ were trained using single nucleus (sn) multiome (snATAC-seq + snRNA-seq) and fluorescence activated nuclei sorted (FANS) Hi-C data from early postnatal to late adulthood human brains^37^ (**Methods**), capturing long-range chromatin interactions and providing an accurate map of regulatory effects on gene expression^38^. Using these E-P interaction maps, we connected COJO SNPs with putative regulatory sequences and credible causal genes (**Extended Data Figure 5A** and Methods) to detect 1,538 E-P interactions across all cell types. 767 unique genes that were putatively associated with 40% of the conditionally independent variants (**Extended Data Figure 5B**; **Supplementary Table 25**) primarily affect E-P interactions within one cell type (**Extended Data Figure 5C**).

Despite the differences in methodologies and the sources of omics data, we found a significant overlap of nominated causal genes predicted by the E-P and TWAS approaches for all major cell types except endothelial cells, for which TWAS reported a markedly lower number of causal genes due to the low frequency of these cells in the training dataset (Bonferroni *P*-value between 7.15×10^−36^-3.30×10^-6^). A total of 94 prioritized protein-coding identifiers were identified across all cell types (**Figure 4D**; **Supplementary Table 25**). In line with prior research, our analysis revealed that these genes are significantly associated with abnormalities in both presynaptic and postsynaptic regions (**Figure 4E**) as they participate in 11 ontological sets in the synaptic organization, covering a variety of specific pathways and functions (**Supplementary Table 26**). For instance, four and three of the prioritized genes encode components of the postsynaptic density membrane (adjusted P-value=0.00175) and the presynaptic active zone membrane (adjusted P-value=0.00175), respectively, highlighting the disruption of several vital processes essential for synaptic transmission.

Among the 94 shared genes, we discovered a connection between schizophrenia COJO GWAS variants rs57022825 and rs17486822 located within a glutamatergic-specific enhancer inside the intronic region of the *NALCN* gene (Sodium Leak Channel Non-Selective Protein; **Figure 4F**). *NALCN* is a major player in determining the influence of extracellular Na+ on a neuron’s basal excitability and its modulation by hormones and neurotransmitters^39^. Although *NALCN* variants have been reported to be in linkage disequilibrium with schizophrenia and bipolar disorder^40,41^, this is the first evidence revealing the cell-specificity of that regulation, providing strong support as it comes from the consensus of two independent methods. Another example is a novel link between an LD buddy of schizophrenia COJO SNP rs6065926, situated within a microglial enhancer, and the cluster of differentiation 40 (CD40) gene (**Figure 4G**). CD40, a member of the tumor necrosis factor receptor (TNFR) superfamily, plays a crucial role in regulating immune responses^42^. Previous research indicated that receptors initiating NF-κB signaling are transcriptionally dysregulated in schizophrenia^43^, but this association had not been established in the context of a common variant affecting microglia-specific regulation of CD40^44,45^. These two examples demonstrate the capability of our model to not only identify causal genes but also, by offering cell-specific contexts, facilitate a better understanding of the mechanisms through which risk loci act.

### Penetrance and pleiotropy of schizophrenia polygenic scores

Understanding the penetrance and pleiotropy of schizophrenia PRS is critical for predicting risk and guiding clinical interventions. We used a “leave-one-out” approach to create an independent PRS constructed from AFR-specific weights, and performed phenome-wide association studies (PheWAS) to explore its relationship with a range of mental and physical health problems.

Across 1,650 unique “phecodes”, PRS were robustly associated with schizophrenia (OR=1.21, 95% CI:[1.18,1.25]; *P*=6.31×10^−44^), bipolar disorder (OR=1.11, 95% CI:[1.09,1.14]; *P*=1.55×10^−16^), major depression (OR=1.05, 95% CI:[1.04,1.07]; *P*<3.89×10^−14^), alcohol-related (OR=1.05, 95% CI:[1.04,1.07]; *P*=1.10×10^−12^) and tobacco use disorders (OR=1.05, 95% CI:[1.03,1.06]; *P*=1.48×10^−10^), and suicidal attempt and ideation (OR=1.09, 95% CI:[1.06,1.12]; *P*=4.10×10^-8^) (**Figure 5A**; **Supplementary Table 27**). We also replicated previously reported associations ^12,47^ between schizophrenia PRS and increased risk of viral hepatitis (OR=1.06, 95% CI:[1.04,1.08]; *P*=4.10×10^-8^), upper respiratory infections (OR=1.04, 95% CI:[1.03,1.06]; *P*=1.77×10^-8^), and dental caries (OR=1.05, 95% CI:[1.03,1.06]; P=1.17×10^−10^), and lower risk of obstructive sleep apnea (OR=0.96, 95% CI:[0.95,0.98]; *P*=3.57×10^-7^) and hypertension (OR=0.96, 95% CI:[0.94,0.98]; *P*=4.88×10^-6^).

**Figure 5.**
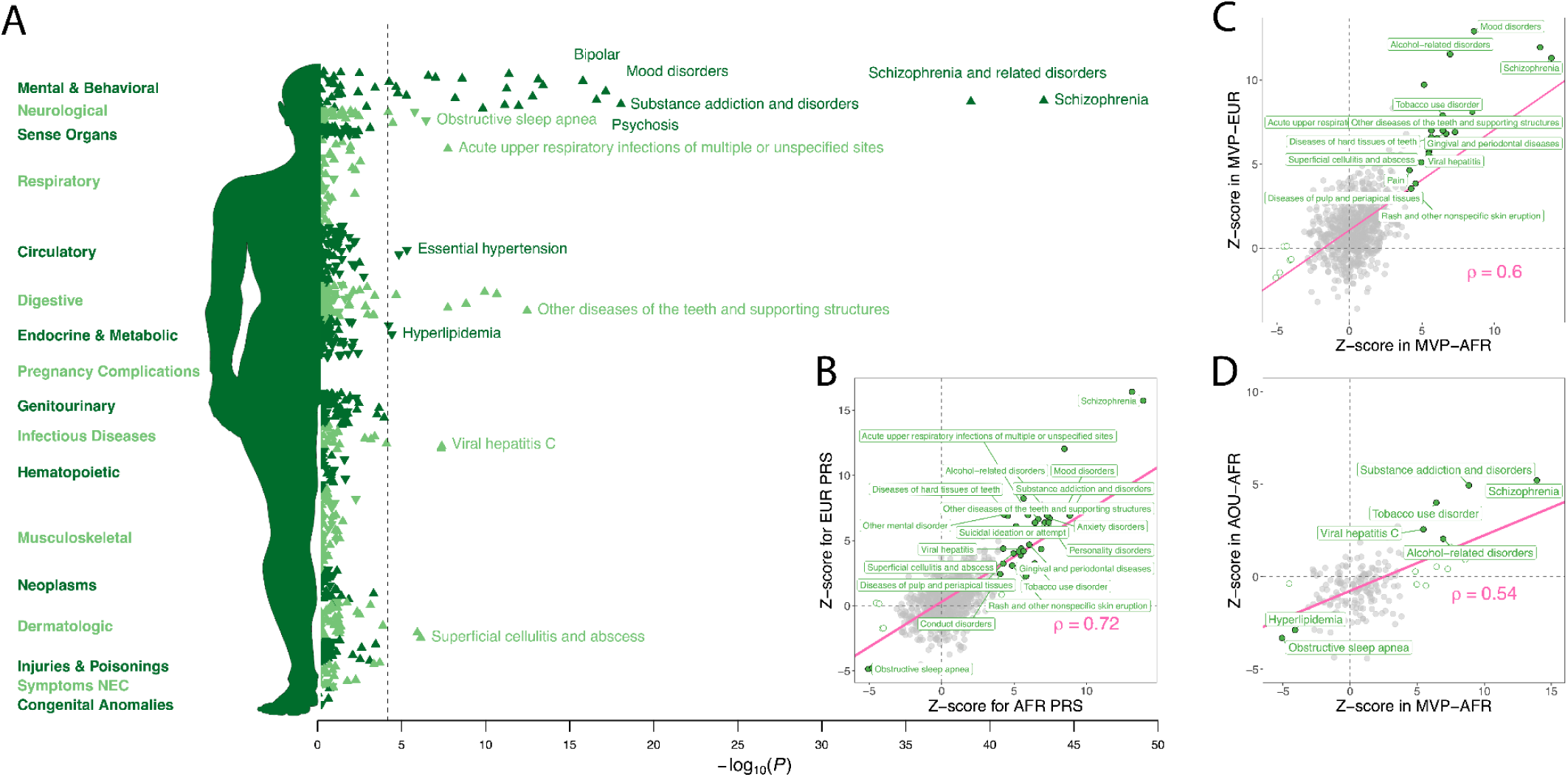
Pleiotropic influences of AFR-derived PRS across diseases. **(A)** PheWAS results for an independent AFR-derived PRS tested against 1,650 disease categories in MVP. The dotted line indicates the Bonferroni adjusted *P*-value threshold for the number of tests. **(B)** Effect sizes for AFR PRS and EUR PRS tested in AFR participants; highlighted points were significant in tests of AFR PRS and labeled if replicated using EUR PRS (*P*<0.05). **(C)** Effect sizes of AFR PRS tested in AFR participants in MVP and AOU; highlighted points were significant in AFR participants, and labeled if replicated in AOU (*P*<0.05). **(D)** Effect sizes of AFR PRS tested in AFR and EUR participants in MVP; highlighted points were significant in AFR participants, and labeled if replicated in EUR (*P*<0.05). Unfilled points (empty circles) represented significant findings in MVP AFR that were not replicated.

We extended this analysis to 49 common laboratory measurements, observing additional significant associations between PRS and increased levels of hematocrit (β=0.020, 95% CI:[0.014,0.026]; *P*=2.98×10^−11^), high density lipoprotein (HDL) cholesterol (β=0.017, 95% CI:[0.011,0.023]; *P*=1.50×10^-7^), and aspartate aminotransferase (AST) (β=0.013, 95% CI:[0.007,0.020]; *P*=6.69×10^-5^) (**Extended Data Figure 7; Supplementary Table 28**).

Overall, we found that findings largely mirrored corresponding results for EUR-trained PRS (**Supplementary Tables 29-30**). We directly compared results based on AFR- and EUR-trained PRS, observing a high degree of correlation (ρ=0.715; *P*=3.01×10^−56^) in effect sizes (**Figure 5B; see Methods**). Notably, the relationship between AFR scores and schizophrenia diagnosis (OR=1.21, 95% CI:[1.18,1.25]; P=6.31×10-44) was slightly attenuated compared to EUR scores (OR=1.32, 95% CI:[1.27,1.36]; P=1.14×10-55), likely reflecting the lower power of our available training data.

Similarly, direct comparison of effect sizes for PheWAS of AFR PRS in AFR versus EUR participants highlighted replicable cross-ancestry associations, and significant overall correlations (ρ=0.6; *P*=1.10×10^−32^) (**Figure 5C**; **Supplementary Tables 31-32**),

Among AFR replication cohorts, EHR data were uniquely available for AOU, presenting an opportunity to replicate our findings across healthcare systems. We found that higher PRS increased the odds of being diagnosed with schizophrenia (OR=1.21, 95% CI:[1.13,1.29]; *P*=1.45×10^-7^) and to a lesser degree, bipolar disorder (OR=1.06 95% CI:[1.01,1.12]; *P*=0.0329); substance addiction (OR=1.11, 95% CI:[1.07,1.15]; *P*=3.47×10^-8^), and tobacco use disorders (OR=1.04, 95% CI:[1.10,1.07]; *P*=5.30×10^-5^). We replicated our previous findings that higher PRS lowered the odds of obstructive sleep apnea (OR=0.93, 95% CI:[0.89,0.97]; *P*=0.0003) and hyperlipidemia (OR=0.95, 95% CI:0.92,0.98]; *P*=0.00385) (**Supplementary Table 33**). Effects estimated in AFR participants in MVP and AOU were significantly correlated (ρ=0.54; *P*=3.17×10^−14^; **Figure 5D**), albeit to a lesser degree than observed for AFR and EUR individuals in MVP. Comparing phenome-wide prevalences across datasets, we observed correlations of 0.94 between AFR and EUR participants, and 0.63 between AFR participants in MVP and AOU (**Supplementary Note**). This finding may reflect different lifetime exposures and patterns of healthcare utilization between military and civilian populations.

## Discussion

This study significantly advances our understanding of schizophrenia’s genetic underpinnings by incorporating data for African ancestry populations. The integration of data from the CSP #572/MVP and AOU Research Programs with four case-control cohorts yielded a sample size of 13,012 affected individuals and 54,266 controls, and increases the sample size of an earlier 2009 study^19^ by a factor of 10, and our 2019^20^ and 2021 studies^7^ by 2.2- and 1.75-fold, respectively. This is the first study to yield independently significant associations in African ancestry populations. Through detailed investigations of heritability, local ancestry, and cell-type enrichment, we underline the shared biological underpinnings of schizophrenia across populations, in spite of heterogeneous genetic effects and pervasive disparities in diagnosis.

The findings reveal both convergent and divergent aspects of the genetic architecture of schizophrenia across different ancestries. While many loci identified in European populations showed directionally consistent effects in individuals of African ancestry, our approach also highlighted loci with potentially population-specific effects. For example, *GRIN2A* harbors both rare coding^48^ and common variants that confer risk for schizophrenia^24^, with distinct associations between other non-coding variation, smoking behaviors^49^, and educational attainment^50^, results that more closely mirror the pattern of association observed in AFR populations.

Differences in allele frequency and patterns of LD have the potential to lower GWAS discovery power, but can also be leveraged via fine-mapping approaches to identify a common, credible set of putatively causal variants. This “trade-off” between power and resolution is best illustrated by the comparatively fewer number of independent SNPs detected in joint analyses of African and European ancestry datasets compared to EUR-EAS and expanded EUR-only analyses, contrasted with the larger reductions in credible set size in joint analyses with AFR. The fine-mapping results, particularly at loci such as *CACNA1I*, demonstrate how leveraging diverse genetic backgrounds can refine the localization of causal variants, potentially accelerating the identification of therapeutic targets.

Focusing on African-like populations uncovered population-specific associations upstream of *PLXNA4*, which encodes a molecule critical for axonal guidance^51–53^, and has been implicated in Alzheimer’s disease^54^, autism^55^, and innate inflammatory responses^56^. Another example is *PMAIP1*, which encodes the proapoptotic molecule, Noxa, a crucial determinant of cell fate in developing oligodendrocytes^57^ that has been shown to be upregulated in the blood of military service members with PTSD^58^ and in frontal brain of mice exposed to olanzapine^59^. We anticipate that further expansions in AFR sample sizes will yield further novel, biologically plausible insights, including allelic heterogeneity and pleiotropic effects. Comparable expansions of EUR sample sizes may yield diminishing returns in terms of locus discovery, as suggested by an independent EUR analysis of 6,579 cases and 155,490 controls from MVP, which recovered three well-established associations at *ZNF804*, *LEMD2*, and *BCL11B*.

Given the selected nature of the population utilizing VHA services, and higher rates of diagnosis among Black and African American individuals, we sought to characterize the common variant genetic architecture of schizophrenia. Directly measured *h*^2^ was virtually indistinguishable between veterans of African and European ancestry^6,7^, which supports the assertion that schizophrenia is no more or less “genetic” in one group versus another. Disparities in the rates of diagnosis could reflect the consequences of structural racism, both in the negative impact of exposure to adverse social conditions^3^, as well implicit biases that label schizophrenia as primarily a “Black disease”^60,61^. Notably, we observed that the stark differences in prevalence of schizophrenia diagnoses appear to have diminished in recent decades (**Supplementary Note**).

The investigation into cell-type-specific genetic architectures revealed significant enrichment of schizophrenia risk loci in excitatory and inhibitory neurons, consistent with previous findings in European populations^24,30,31^. Although aberrant myelination is known to play a role in neuropsychiatric disorders^62^ and the association between OPCs and schizophrenia is well-established^30,31^; however, the higher enrichment of OPCs in African ancestry individuals merits further investigation, as it may uncover novel biological pathways contributing to schizophrenia risk that are more pronounced in, or unique to, these populations.

Our exploration of the polygenic manifestations of currently indexable risk demonstrated resoundingly that AFR-derived scores are robustly transferable across cohorts, settings, and populations. Our PheWAS successfully recapitulated cross-trait associations with mental and physical health conditions, including both risk-increasing and apparently protective effects, and many which remained significant when limiting our analyses to individuals without a formal psychiatric diagnosis.

Despite nearly doubling the sample size of prior AFR GWAS^7,20^, we were still underpowered relative to contemporary EUR^24^ and EAS^23^ studies. Pointedly, our analysis is on a par with the discovery phase of a 2013 study that yielded genome-wide significant associations in 12 genomic regions^63^. It is important to note that the assembled study cohorts all originated in the United States, and therefore represent a fraction of the vast genomic (and experiential) diversity present in African diaspora populations. By definition, heritability is a population-specific phenomenon, and forthcoming research initiatives across continental Africa and the Caribbean^64,65^ have tremendous potential to accelerate genetic discovery and epidemiological insights. Lastly, though schizophrenia is a highly heritable disorder, there is considerable evidence for the role of social conditions, especially those related to structural racism^2,3^. Approaches that integrate social and biological risk factors are important for improving our understanding of the etiology of schizophrenia.

In conclusion, this study not only contributes valuable data to the field of psychiatric genetics but also highlights the critical importance of including diverse populations in genetic research. Large-scale genome sequencing endeavors should prioritize genetically diverse and otherwise underrepresented populations in order to fully realize their potential for novel discoveries^64,66^ and realize biological insights with equitable public health relevance to patients and communities worldwide, paving the way for advancements in precision medicine that are inclusive and beneficial for all populations.

## Online Methods

### Ethics/study approval

This study was approved by the VA Central Institutional Review Board (IRB), and participating studies received approval from their respective IRBs. All participants provided written informed consent.

### Study Participants

Details of study ascertainment and assessment for the CSP/MVP, All of Us, COGS, GPC, PAARTNERS, and MGS are summarized in the **Supplementary Note**, and have otherwise been described elsewhere^7,12,14–16,18–21,67,68^

### Genome-wide association studies, replication, and meta-analysis

We performed primary GWAS of schizophrenia in CSP/MVP, COGS, MGS, and PAARTNERS using imputed allelic dosages and logistic regression, as implemented in PLINK2, adjusting for age, sex, and 10 ancestry principal components (PCs).

For AOU, we performed logistic regression as implemented in HAIL using whole-genome sequencing data (release 7), and adjusting for age, sex, and 10 ancestry PCs.

We meta-analyzed AFR cohorts using inverse variance weighted meta-analysis (“fixed effects”), and combined results across ancestries using Han and Eskin’s random effects model with heterogeneity (RE2)^69^. Both models are implemented in the METASOFT software package.

### Heritability estimation

We directly estimated heritability (SNP-*h*^2^) from imputed genotypes using genome-based restricted maximum likelihood (GREML)^70^ as implemented in the genome-wide complex trait analysis (GCTA) software^70,71^. We included the same GWAS covariates and used the [--grm-cutoff .05] flag to restrict analyses to approximately unrelated individuals. To enhance comparability of results, we downsampled the CSP/MVP data to include 6,000 cases and 18,000 screened controls of each ancestry (AFR and EUR). We estimated minor allele frequencies within each ancestry, and the absolute frequency difference (AFD)^29^, and partitioned SNPs into bins based on ad hoc MAF thresholds or AFD quintiles, followed by partitioned heritability analysis^70^. For ease of interpretability in the context of the published literature, we performed liability scale transformations assuming a 1% lifetime prevalence of schizophrenia.

### Fine-mapping and credible sets

We used the Sum of Single Effects model (SuSiE)^72,73^ as implemented in the SuSiE-R package, to estimate causal credible SNP sets from GWAS meta-analysis results. First, we defined associated intervals around conditionally independent SNPs, collapsing all COJO variants within 250 kb windows. We calculated reference LD information for “super populations” in the 1000 Genomes Population Phase 3 data (i.e. EUR, AFR, EAS). Briefly, for each ancestral population represented in a given GWAS meta-analysis, we applied the following quality control steps: 1) we removed strand-ambiguous SNPs, multi-allelic SNPs, and SNPs with MAF less than 0.1%; 2) we retained only common SNPs across these ancestries and present the corresponding LD reference data. Next, we estimated credible sets at each associated interval, defined as the minimum number of ranked variants with cumulative posterior inclusion probability (PIP) greater than or equal to 0.95. We estimated credible sets based on EUR results alone, comparing these to corresponding sets derived from cross-ancestry GWAS findings. We evaluated improvements in fine-mapping on the basis of the size (the number of SNPs included in the set) and interval length (the total kb encompassed by each set). We used a probability threshold of 0.95 to define credible sets, and tentatively classified as causal any SNP with a posterior inclusion probability (PIP) >0.5.

We extended these analyses to the direct comparisons of ancestry-specific GWAS results using a recent extension of SuSiE, multi-ancestry sum of the single effects model (MESuSiE)^74^. Briefly, MESuSiE models ancestry-specific information for two populations (e.g., EUR-AFR, EUR-EAS, AFR-EAS) to enhance the precision and resolution of statistical fine-mapping. Like SuSiE-R, this method takes summary statistics as input, considers the varied LD patterns present in distinct ancestries, explicitly models shared and ancestry-specific causal effects, and employs a scalable variational inference algorithm for computation. SNPs in the resulting 95% credible sets had a non-zero effect in at least one population, and are classified as either “shared” or “ancestry-specific” depending on their modeled contribution to the credible set.

We also considered SuSiE-X^75^, another extension of the SuSiE framework, which has been shown to enhance precision of cross-population fine-mapping while maintaining well-calibrated false positive rates, and retaining the capability to identify population-specific causal variants. We followed the same steps as described above for processing ancestry-specific GWAS and LD information.

### Local ancestry inference

We employed RFMix2^76^ to deconvolve local ancestry among admixed AA individuals, implementing a local ancestry adjusted model analogous to the tractor method^25^. Specifically, we simultaneously modeled haploid dosages for AFR- and EUR-derived tracts, along with the number of AFR haplotypes carried by an individual at a locus (i.e. “hapcount”) using the local covariate functionality of PLINK2^77^.

To better understand how admixture can impact the “portability” of schizophrenia findings, we compared conventional GWAS results for AFR and EUR (i.e. with global PC adjustment) with estimates based on deconvolved haplotypes in admixed AFR individuals. Across 270 replicated schizophrenia associations, 65% and 90% showed the same direction of effect in AFR (*P*=1×10^-7^) and EUR participants in MVP. When considering only AFR tracts in AFR participants, only 58% (*P*=0.007) of SNPs showed directionally consistent effects, compared to 69% (*P*=3.4×10^−10^) based on EUR tracts.

### Cell-type enrichment at single cell resolution

We used the single-cell Disease-Relevance Scoring (scDRS)^33^ methodology to evaluate the aggregate expression of potential disease-associated genes obtained from EUR and AFR summary statistics utilizing Multi-marker Analysis of GenoMic Annotation (MAGMA)^78^. Each putative disease gene was weighted by its MAGMA *Z*-score from GWAS and inversely weighted by its gene-specific technical noise level in single-cell data obtained from an extensive dataset of 6.3 million nuclei generated from postmortem human dorsolateral prefrontal cortex of 1,494 donors. This process generated cell-specific raw disease scores. Additionally, we computed 1,000 sets of cell-specific raw control scores from matched control gene sets, ensuring similarity in gene set size, mean expression, and expression variance with the putative disease genes. Subsequently, we normalized both the raw disease scores and raw control scores for each cell, yielding normalized disease scores and normalized control scores. The computation of these scores utilized the default settings of the scDRS compute-score function. For downstream analysis, we conducted cell type-level assessments to link broad cell types to disease and explore heterogeneity in disease association across cells within each broad cell type. This was achieved using the scDRS *perform-downstream()* function with default settings. To address multiple testing, false discovery rate (FDR) was calculated using the Benjamini-Hochberg method.

### TWAS concordance

To perform TWAS analysis, we utilized brain single nucleus (sn) transcriptomic imputation models from the PsychAD cohort. S-PrediXcan^79^ was used to perform ancestry-matched TWASs for schizophrenia GWASs. Consequently, TWAS z-scores across all cell-type/gene pairs were normalized (mean = 0; SD=1) within each ancestry to more easily compare results across ancestries. To assess concordance, only common cell-type/gene pairs were retained. Genes were ranked by meta-analysis p-value (performed on scaled z-scores). Correlation was calculated amongst all common genes in each cell type, and sequentially, genes were restricted to increasingly smaller sets (top 1,000 genes, 990 genes, etc.) to construct correlation trajectories. Only well-represented class-level cell types were retained for visualization.

### Prediction of enhancer-gene interactions

To study E-P interactions affected by schizophrenia GWAS variants, we leveraged cell-type-specific E-P maps from a multi-omics dataset (joint snATAC-seq & snRNA-seq and cell-specific Hi-C) of developing brains using activity-by-contact (ABC) model^36,37^ (**Extended Data Figure 6A**). In line with authors’ instructions^36,38^, we filtered out E-P interactions that (i) had an ABC score < 0.015, (ii) involved ubiquitously expressed genes or genes on the Y chromosome, and (iii) included genes not expressed in major brain cell types. To further refine our focus to schizophrenia GWAS, we decided to keep only E-P links (i) overlapping conditionally independent SNPs (peaks participating in E-P links were extended by 100bp on both sides to increase the overlap), (ii) LD buddies of conditionally independent SNPs (R2 ≥ 0.8), or (iii) SNPs within 95% credible set and PPI > 0.5 (model AFR+EUR+EAS), resulting in 34,436 E-P links. To limit the number of causal genes per conditionally independent SNPs, we applied the ABC-MAX approach^38^, allowing only one E-P per conditionally independent SNP, resulting in 1,538 E-P links (**Extended Data Figure 6B**).

### PRS Profiling

We constructed PGS from published and current GWAS results (the “training” datasets), testing individual-level scores for association with case-control status in MVP or replication cohorts (the “target” dataset). We applied a recently-developed Bayesian framework that applies continuous shrinkage to test statistics, PRS-CSx^27^, using HapMap 3 SNPs and 1000 Genomes Project LD reference panel (EUR and AFR). We also constructed scores based on genome-wide significant SNPs, following LD-based clumping in the appropriate KGP3 population (r^2^>0.1; 500kb window). Scores were constructed by summing the number of tested alleles weighted by their effect estimates (e.g., the log of the allelic odds ratio). We tested for case-control differences by logistic regression using age, sex, and the first six ancestry principal components (PCs).

### PheWAS and LabWAS

We employed phenome-wide association studies (PheWAS) to explore the relationships between neuropsychiatric PRSs and “phecodes” representing groupings of related ICD-9/10 billing codes^80^.When testing individual phecodes, we required cases and controls to have ≥2 and zero codes, respectively. We applied logistic regression to test scaled PRSs (mean=0; SD=1) for association with phecodes within ancestry groups, covarying for age, age^2^, gender, and six ancestry principal components (PCs). We performed LabWAS across mean, median, maximum, and minimum normalized values in individuals with at least two observed measurements using linear regression and the same covariates as in PheWAS analyses.

We performed a series of sensitivity analyses, co-varying for selected diagnoses or treatment with antipsychotics, mood-stabilizers, and antidepressants, or removing individuals with any lifetime diagnosis of psychotic, mood, or substance disorders.

We compared effect sizes across PheWAS results sets using an pseudo-independent set of phecodes, based on the most significant among ontologically nested terms. We calculated Pearson’s correlations based on standardized effect sizes (i.e., *Z*-scores).

## Reporting summary

Additional information about our study design is available in the Nature Research Reporting Summary linked to this paper.

## Data availability

All data produced in the present study are available upon reasonable request to the authors

## Code availability

We utilized second generation PLINK for primary quality control and descriptive analyses (https://www.cog-genomics.org/plink2), For haplotype phasing and genotype imputation, we used SHAPEIT4 (https://odelaneau.github.io/shapeit4/) and Minimac4 (https://github.com/statgen/Minimac4), respectively. For local ancestry inference and defining ancestry tracts, we utilized RFMix2 (https://github.com/slowkoni/rfmix). Conditional and joint analysis (COJO) and heritability estimation utilized GCTA (https://cnsgenomics.com/software/gcta). Meta-analyses were conducted using METASOFT (https://www.zarlab.xyz/tag/metasoft/). For statistical fine mapping, we used SuSiE-R (https://github.com/stephenslab/susieR), SuSiE-X (https://github.com/getian107/SuSiEx), and MESuSiE (https://github.com/borangao/MESuSiE). We estimated gene-level test statistics using MAGMA (https://cncr.nl/research/magma/) and tested for cell-type specific enrichment using scDRS (https://github.com/martinjzhang/scDRS). We used S-PrediXcan (https://github.com/hakyimlab/MetaXcan) to directly impute TWAS results from GWAS summary statistics. To characterize enhancer-gene links, we adopted the ABC-MAX framework (https://github.com/EngreitzLab/ABC-Max-pipeline). For PRS, we used PRS-CSx to integrate GWAS summary statistics from multiple populations and estimate posterior SNP effects via continuous shrinkage (https://github.com/getian107/PRScsx).

## ACKNOWLEDGEMENTS

This research was supported by the Department of Veterans Affairs Cooperative Studies Program (CSP) #572 and the Million Veteran Program (MVP-000, MVP-006 and MVP-076). The Million Veteran Program is supported by the Office of Research and Development, Department of Veterans Affairs. The contents do not represent the views of the U.S. Department of Veterans Affairs or the United States Government. Complete acknowledgements for CSP #572 and MVP are given in the Supplementary Materials.

This study was supported by the Veterans Affairs Merit grants: BX004189 (to P.R.). This work was supported by the National Institutes of Health (NIH): R01MH125246 (to P.R. and T.B.), K08MH122911 (to G.V.), R01AG078657 (to G.V.), R01MH104964 (to T.B., A.H.F., M.T.P., C.N.P.), R01AG067025 (to P.R.), U01MH116442 (to P.R.), and U24AG087563 (to P.R.).

The GPC was supported by grants R01 MH085548 and R01 MH104964 from the National Institute of Mental Health (NIMH), and genotyping of samples was provided by the Stanley Center for Psychiatric Research at Broad Institute.

COGS was supported by grants R01 MH065571, R01 MH065588, R01 MH065562, R01 MH065707, R01 MH065554, R01 MH065578, R01 MH065558, R01 MH86135, and R01 MH094320 from the National Institute of Mental Health.

Funding support for the Genome-Wide Association of Schizophrenia Study was provided by the NIMH (R01 MH67257, R01 MH59588, R01 MH59571, R01 MH59565, R01 MH59587, R01 MH60870, R01 MH59566, R01 MH59586, R01 MH61675, R01 MH60879, R01 MH81800, U01 MH46276, U01 MH46289, U01 MH46318, U01 MH79469, and U01 MH79470) and the genotyping of samples was provided through the Genetic Association Information Network (GAIN).

Dr. Harvey has served as a consultant to multiple pharmaceutical companies and device manufacturers on phase 2 or 3 development; this consulting work has been determined to be unrelated to the content of the paper. No other authors report any relevant conflicts of interest.

## EXTENDED TABLES AND FIGURES

**Extended Data Table 1.** Sample sizes and characteristics of primary and replication datasets. For each *Study*, grouped by broad ancestry group (*Pop*), *Source* indicates whether genotypes, sequencing data, or summary statistics were directly accessed, *Dx* gives the clinical source for schizophrenia diagnoses, and Screened indicates whether control participants were screened for relevant psychiatric conditions.

| Pop. | Study | Source | Cases |  | Controls |  | *N*<sub>combined</sub> |
| --- | --- | --- | --- | --- | --- | --- | --- |
|  |  |  | Dx | *N* | Screened? | *N* |  |
| AFR | CSP #572 | Genotype | SCID | 1,920 | Yes | 6,000 | 7,920 |
|  | MVP* | Genotype | ICD-9/10 | 4,488 | Yes | 14,065 | 18,553 |
|  | All of Us* | WGS | ICD-9/10 | 1,065 | Yes | 27,866 | 28,931 |
|  | GPC | Genotype | DI-PAD | 3,506 | Yes | 3,849 | 7,355 |
|  | COGS | Genotype | DIGS | 498 | Yes | 405 | 903 |
|  | MGS | Genotype | DIGS | 994 | Yes | 1,277 | 2,271 |
|  | PAARTNERS | Genotype | DIGS | 541 | No | 804 | 1,345 |
|  |  |  |  | **13,012** |  | **54,266** | **67,278** |
| EUR | CSP #572 | Genotype | SCID | 1,236 | Yes | 29,200 | 30,436 |
|  | MVP* | Genotype | ICD-9/10 | 5,343 | Yes | 126,290 | 131,633 |
|  | PGC3 | Summary statistics | Multiple | 53,386 | Both | 77,258 | 130,644 |
|  |  |  |  | **59,965** |  | **232,748** | **292,713** |
| EAS | PGC3 | Summary statistics | Multiple | 14,004 | Both | 16,757 | 30,761 |
| **Totals** |  |  |  | **86,981** |  | **303,771** | **390,752** |

Abbreviations: Cooperative Studies Program (CSP); Genomic Psychiatry Cohort (GPC); Consortium on the Genomics of schizophrenia (COGS); Molecular Genetics of schizophrenia (MGS); Project Among African-Americans to Explore Risks for schizophrenia (PAARTNERS); Structured Clinical Interview for DSM-IV (SCID); International Statistical Classification of Diseases (ICD); Diagnostic Interview for Genetics Studies (DIGS); Whole-genome sequencing data (WGS).

**Extended Data Table 2.** Genome-wide significant (*P*<5×10^-8^) associations in AFR GWAS of schizophrenia. For conditionally independent SNPs, the chromosome and genomic coordinates (GRCh38), *Tested* or effect allele, and population frequency in the gnomAD browser (v4.1.0) are displayed. *Beta* and *Std Error* are the allelic effect size and its standard error; *P*_logit_ is its statistical significance from the basic logistic regression model; *P*_FE_ is its significance based inverse variance weighted (fixed effects); *P*_RE2_ is its significance based on Han and Eskin’s random effects heterogeneity model, and *P*_het_ is the significance of Cochran’s heterogeneity test.

| SNP | Chr | GRCh38 position | Tested | Freq | Phase 1 |  |  | Phase 2 |  |  |  |  |
| --- | --- | --- | --- | --- | --- | --- | --- | --- | --- | --- | --- | --- |
| | | | | | Beta | Std Error | $P_{\text{logit}}$ | Beta | Std Error | $P_{\text{FE}}$ | $P_{\text{RE2}}$ | $P_{\text{het}}$ |
| chr3:112442573:A:C | 3 | 112,442,573 | A | 0.352 | -0.122 | 0.022 | **$3.79 \times 10^{-8}$** | -0.061 | 0.016 | $2.13 \times 10^{-4}$ | $3.47 \times 10^{-6}$ | $6.02 \times 10^{-3}$ |
| chr3:114552436:T:A | 3 | 114,552,436 | A | 0.141 | 0.178 | 0.031 | **$1.11 \times 10^{-8}$** | 0.100 | 0.023 | $1.03 \times 10^{-5}$ | $3.27 \times 10^{-7}$ | $5.50 \times 10^{-3}$ |
| chr7:132080559:C:T | 7 | 132,080,559 | T | 0.158 | 0.153 | 0.029 | $1.36 \times 10^{-7}$ | 0.120 | 0.021 | **$1.91 \times 10^{-8}$** | $3.34 \times 10^{-8}$ | 0.202 |
| chr8:72154435:G:A | 8 | 72,154,435 | A | 0.156 | -0.126 | 0.030 | $2.90 \times 10^{-5}$ | -0.123 | 0.022 | **$2.26 \times 10^{-8}$** | $3.99 \times 10^{-8}$ | 0.899 |
| chr16:10063406:C:A | 16 | 10,063,406 | A | 0.383 | 0.121 | 0.022 | **$1.84 \times 10^{-8}$** | 0.059 | 0.016 | $1.84 \times 10^{-4}$ | $2.51 \times 10^{-6}$ | $3.86 \times 10^{-3}$ |
| chr17:6264499:G:A | 17 | 6,264,499 | A | 0.010 | 0.546 | 0.097 | **$1.63 \times 10^{-8}$** | . | . | . | . | . |
| chr17:48458465:TCA:T | 17 | 48,458,465 | T | 0.009 | 0.641 | 0.108 | **$2.48 \times 10^{-9}$** | . | . | . | . | . |
| chr18:59852986:G:A | 18 | 59,852,986 | A | 0.272 | -0.138 | 0.025 | **$2.53 \times 10^{-8}$** | -0.102 | 0.018 | **$2.02 \times 10^{-8}$** | $2.71 \times 10^{-8}$ | 0.349 |

**Extended Data Fig. 1.**
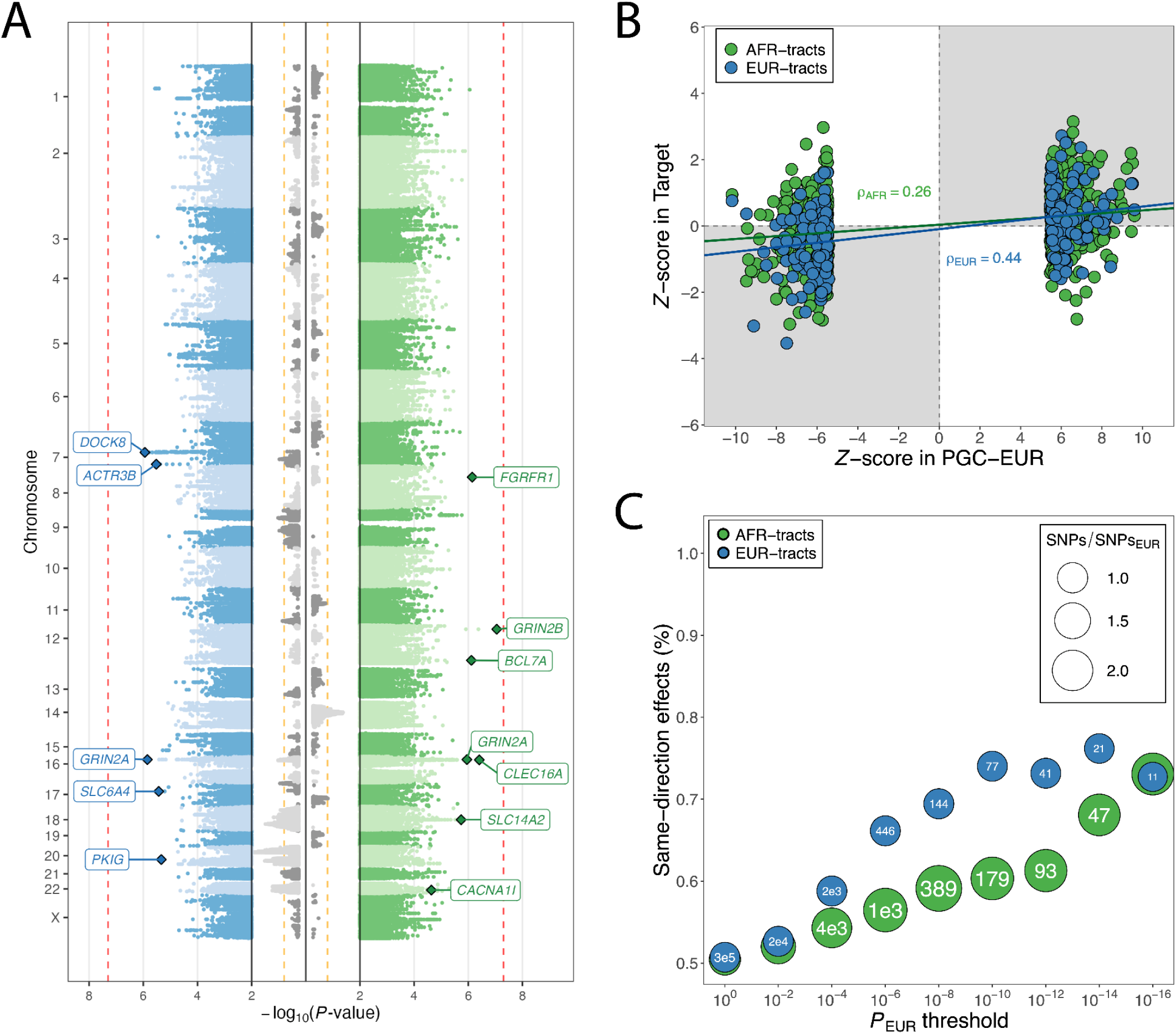
Tract-based GWAS results and cross-ancestry concordance. **A**) Miami plot displaying tract-level association results from the joint analysis of EUR tracts (*left*), AFR tracts (*right*), and haplotype count (*center*). Selected, suggestive findings (*P*<10^-5^) within 100kb of a protein-coding gene are highlighted. In the center panel, the haplotype count results are plotted directionally to indicate which ancestral haplotype is risk-increasing. **B)** Standardized effect estimates for AFR- and EUR-tracts at genome-wide significant SNPs in PGC-EUR, with regression lines; panels shaded in gray contain findings with directionally concordant effects. **C)** Comparison of directional concordance of AFR- and EUR-tract based results with PGC-EUR findings.

**Extended Data Fig. 2.**
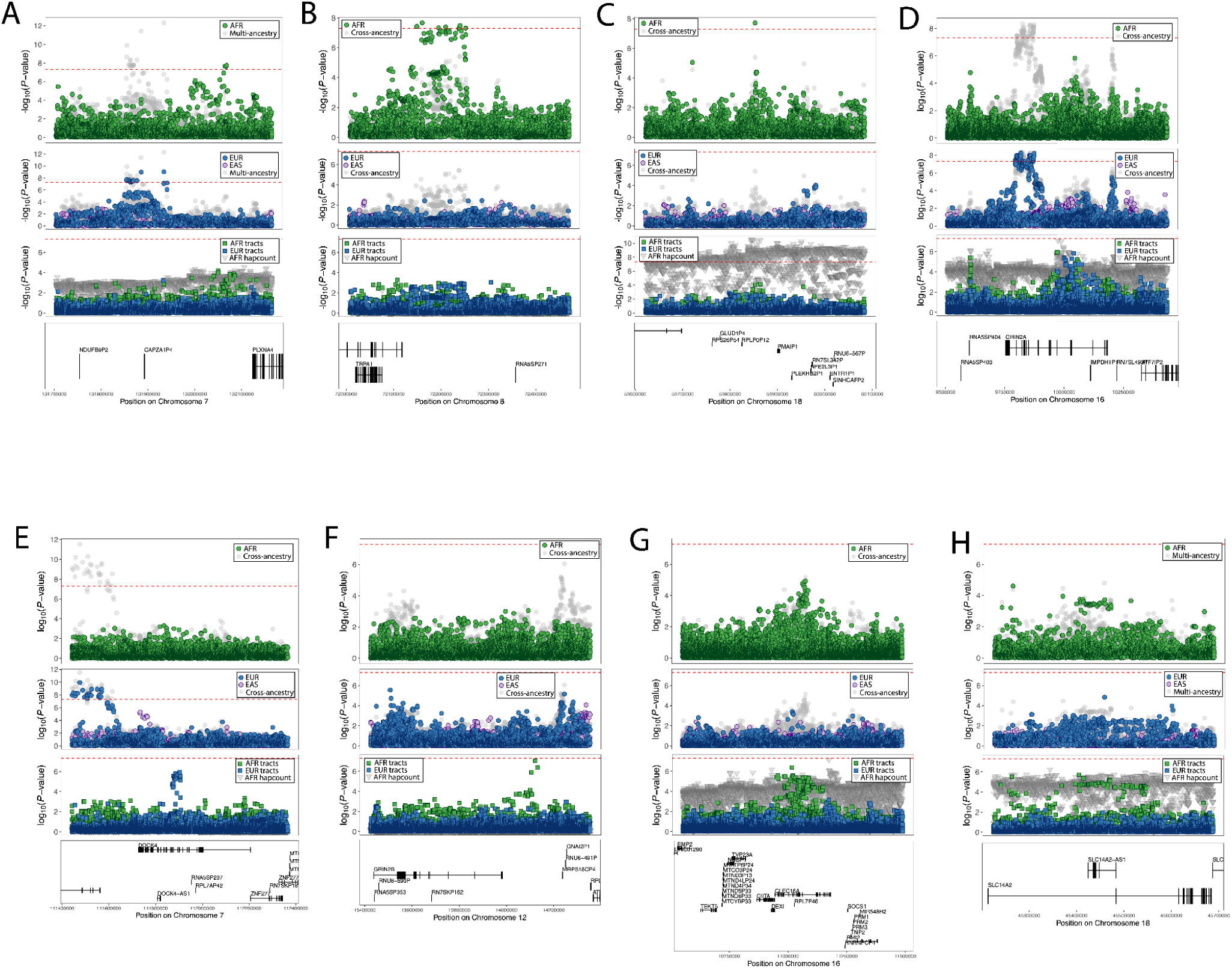
Regional association plots displaying distinct ancestry-specific evidence of association. In each plot, the top panel displays AFR and multi-ancestry meta-analysis results; the middle panels display EUR and EAS results; and the third panel displays results for deconvolved AFR and EUR tracts and AFR haplotype count; genic context is displayed underneath.

**Extended Data Fig. 3.**
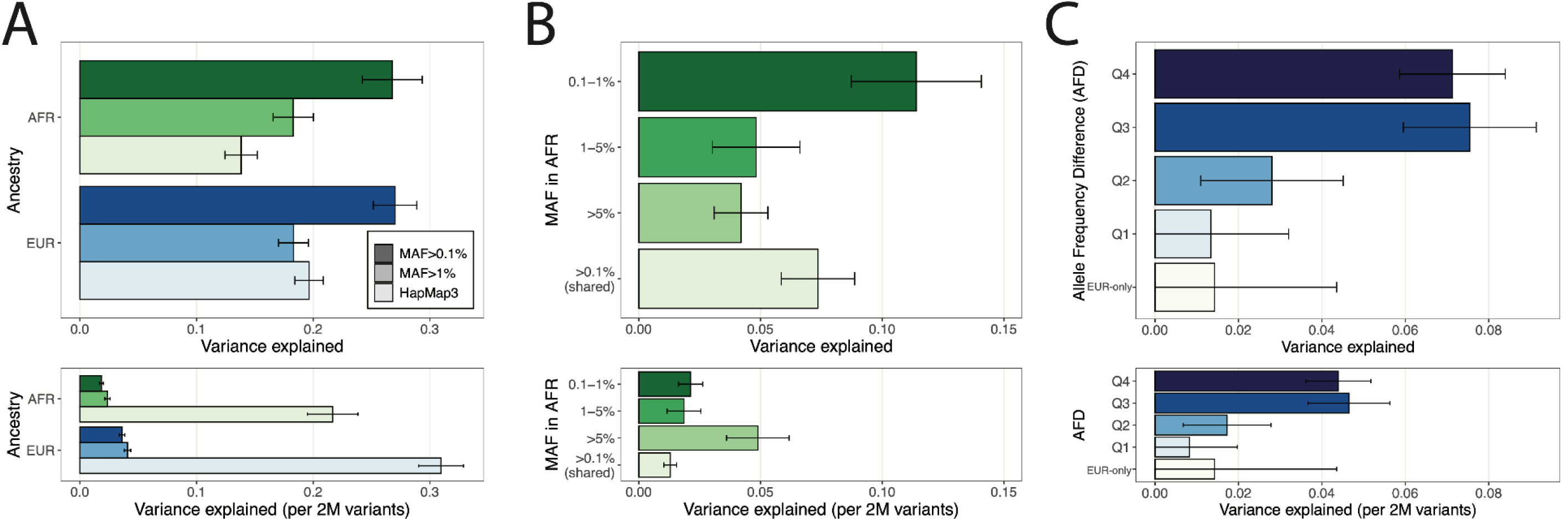
SNP-based heritability of schizophrenia in AFR and EUR individuals. **A)** Estimates of SNP-based *h*^2^ based on progressively more inclusive SNP criteria, comparing HapMap3 SNPs with SNPs with population-specific MAF greater than 1% or 0.1%. **B)** In AFR individuals, partitioned SNP-*h*^2^ for SNPs shared with EUR populations and those unique to the AFR dataset, binned by AFR-specific MAF. **C)** In EUR individuals, partitioned SNP-*h*^2^ for SNPs present only in EUR and those shared with AFR populations, binned by absolute frequency difference (AFD) between EUR and AFR. In each figure, the upper panel displays the total SNP-*h*^2^ and the lower panel displays SNP-*h*^2^ scaled by the number of SNPs used to calculate the genetic related matrix (GRM), respectively. SNP-*h*^2^ is reported on the liability scale, based on an expected population prevalence of 1%, to enhance interpretability with respect to the published literature; observed scale estimates are reported in Supplementary Tables 21-24.

**Extended Data Fig. 4.**
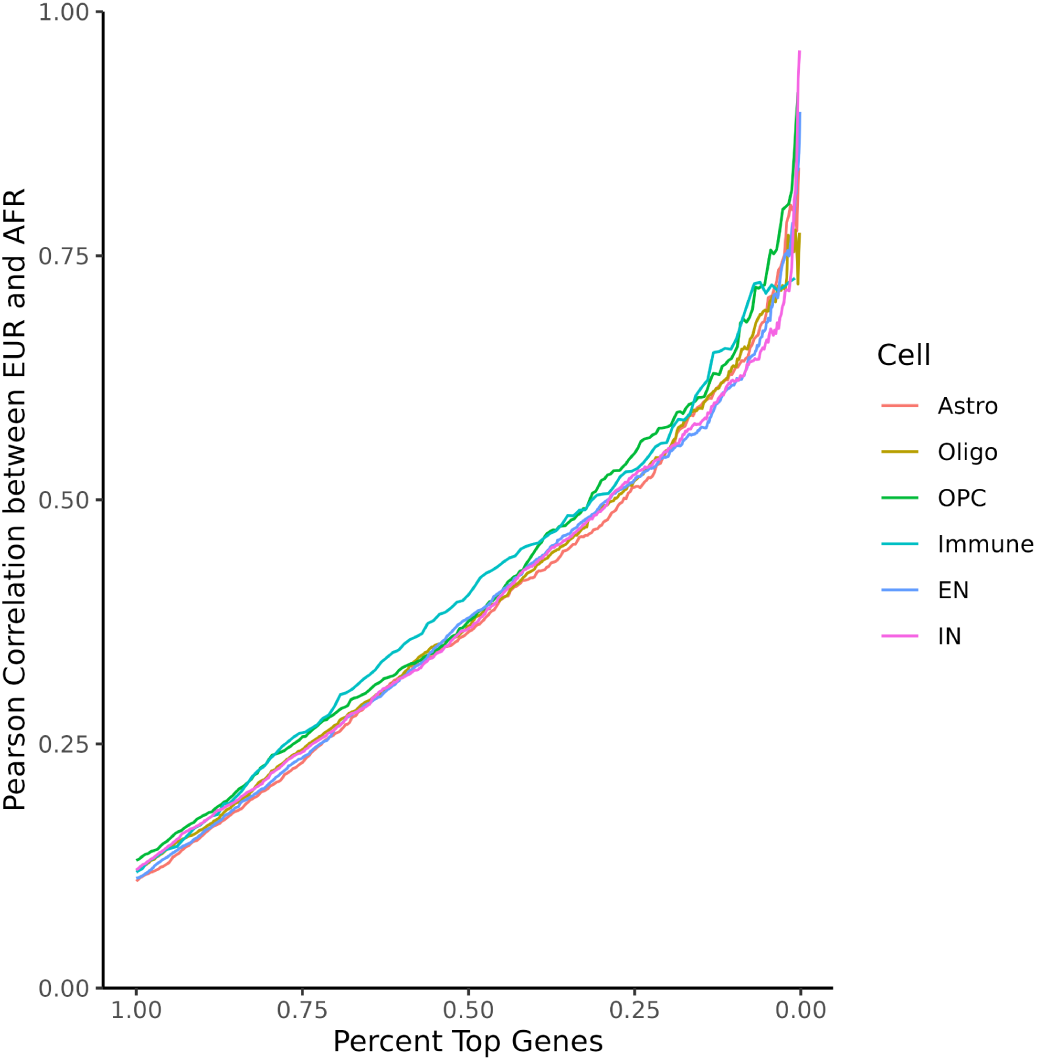
Line represents the pearson correlation between EUR and AFR schizophrenia TWAS amongst the top X% (x-axis) ranked TWAS genes (ranked by fixed-effect meta-analysis performed on scaled z-scores from each ancestry TWAS). Color represents the cell-type in which each TWAS was performed. Genes are limited to those which are imputable within both ancestries in each cell-type. Imputable genes are considered passing cross-validation R^2^ (R^2^_CV_) ≥ 0.01, p_CV_ ≤ 0.05 and SNPs in model > 0. R^2^_CV_ and p_CV_ values are prediction performance R^2^ and prediction performance p-value from the “predict db” software.

**Extended Data Fig. 5.**
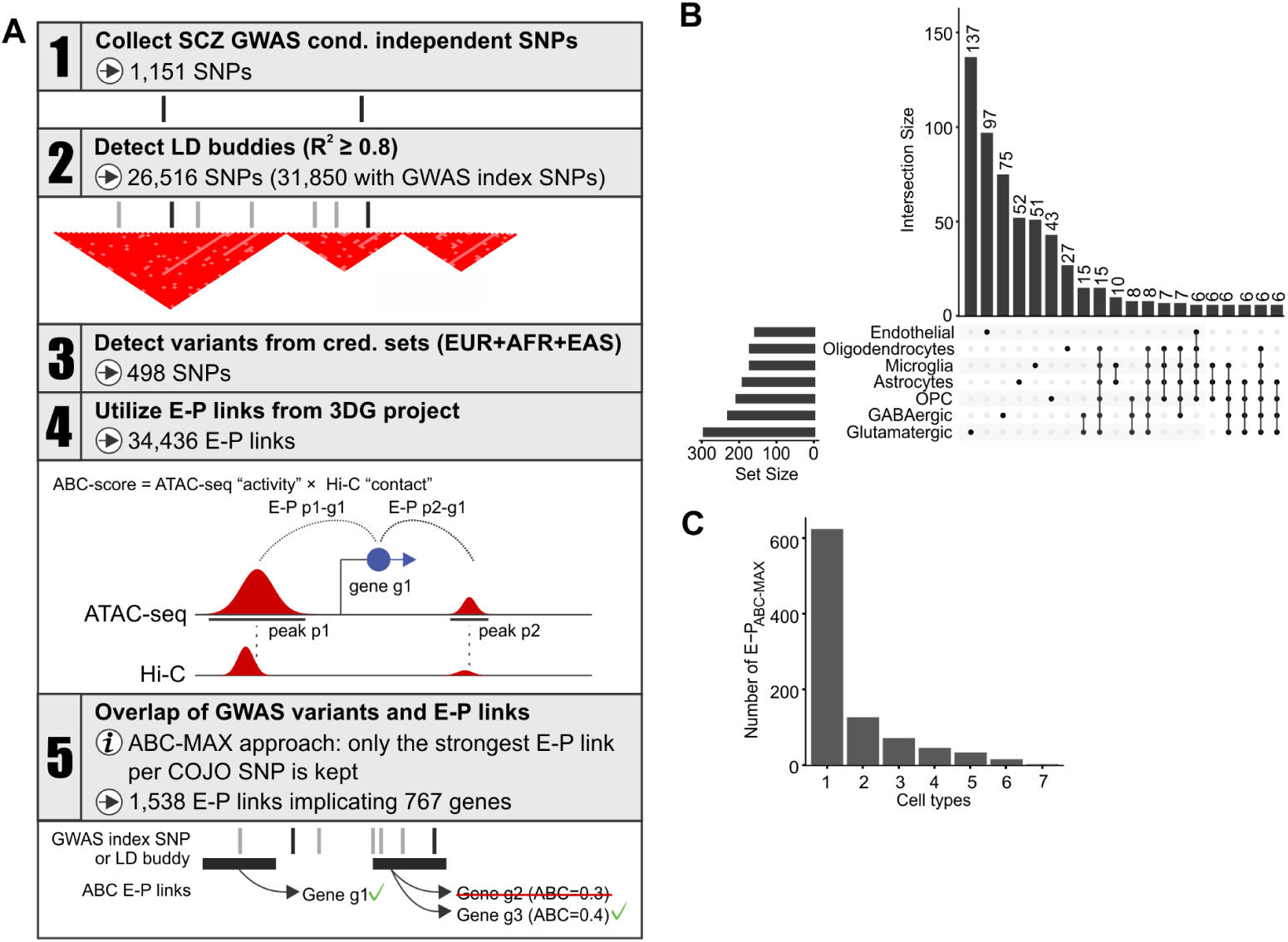
**A)** Schematic of the overall strategy to connect risk variants associated with schizophrenia to their causal genes. **B)** Numbers of nominated causal genes and their overlaps among major brain cell types (minimum intersection size for plotting: 6). **C)** Histogram of distribution of E-P_ABC_ across major brain cell types.

**Extended Data Fig. 6.**
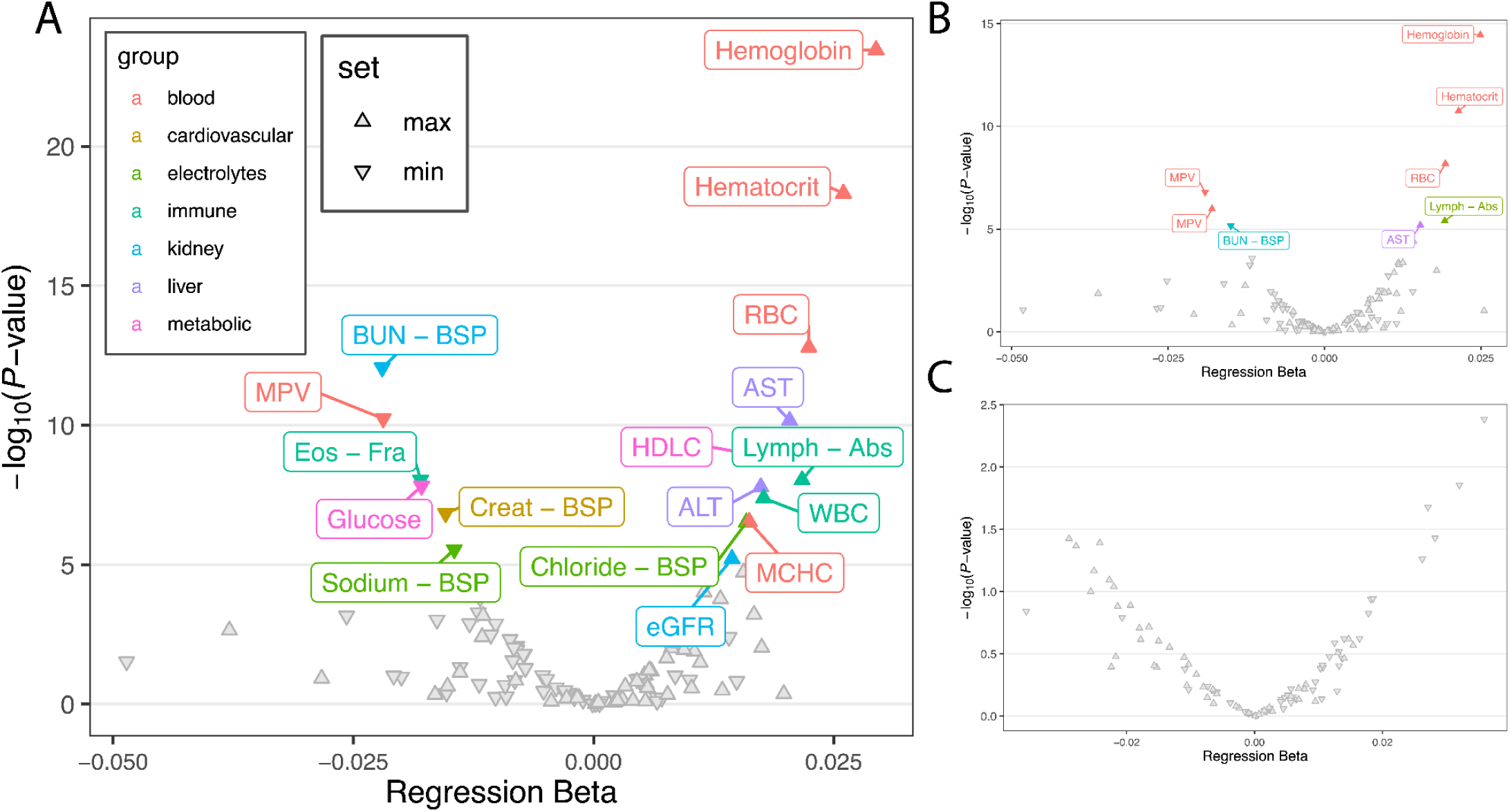
Association of AFR-trained PRS with common laboratory tests (LabWAS). **A)** Volcano plot displaying tested associations in all participants. Highlighted points were significant following Bonferroni adjustment for multiple-testing. Upwards and downwards facing triangles indicate if an observed association is with the highest or lowest observed values across participants’ EHRs. **B)** Corresponding results based on AFR screened controls. **C)** Corresponding results based on case-only analysis.

## CONSORTIA AUTHORS

### Cooperative Studies Program (CSP) #572

Mihaela Aslan ^PC,EC,1^, M. Antonelli ^PC^, John Concato ^PC,EC,1^, F. Cunningham ^PC^, M. Gaziano ^PC,EC,2^, Theresa Gleason ^PC,3^, Philip D. Harvey ^PC,EC,4^, Grant D. Huang ^PC,3^, Thomas R. Kosten ^PC,EC,6^, Alan C. Swann ^6^, P. Miller ^PC,1^, Timothy J. O’Leary ^PC,3^, Ronald Przygodzki ^PC,3^, Pamela Sklar ^PC,EC,5^, Hongyu Zhao ^PC,EC,1^, Ayman Fanous ^EC,20,7^, Shrikant Mane ^1^, M. Corsey ^5^, L. Zaluda ^5^, J. Johnson ^4^, D. Cavaliere ^11^, V. Jeanpaul ^1^, Alysia Maffucci ^1^, L. Mancini ^1^, P. Peduzzi ^PC,1^, J. Deen ^2^, G. Muldoon ^2^, S. Whitbourne ^2^, L. Adamson ^8^, L. Calais ^8^, G. Fuldauer ^8^, R. Kushner ^8^, G. Toney ^8^, M. Lackey ^8^, A. Mank ^8^, N. Mahdavi ^8^, G. Villarreal ^8^, E. C. Muly ^9^, F. Amin ^9^, M. Dent ^9^, J. Wold ^9^, B. Fischer ^10^, A. Elliott ^10^, C. Felix ^10^, G. Gill ^10^, P. E. Parker ^11^, C. Logan ^11^, J. McAlpine ^11^, Lynn E. DeLisi ^12^, S. G. Reece ^12^, M. B. Hammer ^12^, M. de Asis ^PC,12^, D. Agbor-Tabie ^13^, W. Goodson ^13^, M.S. Bauer ^PC,14^, Mary Brophy ^PC,EC,14^, M. Aslam ^14^, M. Grainger ^14^, Neil Richtand ^14^, Alexander Rybalsky ^14^, R. Al Jurdi ^6^, E. Boeckman ^6^, T. Natividad ^6^, D. Smith ^6^, M. Stewart ^6^, S. Torres ^6^, Z. Zhao ^6^, A. Mayeda ^15^, A. Green ^15^, J. Hofstetter ^15^, S. Ngombu ^15^, M. K. Scott ^15^, A. Strasburger ^15^, J. Sumner ^15^, G. Paschall ^16^, J. Mucciarelli ^16^, R. Owen ^16^, S. Theus ^16^, D. Tompkins ^16^, S. G. Potkin ^17^, C. Reist ^17^, M. Novin ^17^, S. Khalaghizadeh ^17^, R. Douyon ^4^, N. Kumar ^4^, B. Martinez ^4^, S. R. Sponheim ^18^, T. L. Bender ^18^, H. L. Lucas ^18^, A. M. Lyon ^18^, M. P. Marggraf ^18^, L. H. Sorensen ^18^, C. R. Surerus ^18^, C. Sison ^19^, J. Amato ^19^, D. R. Johnson ^19^, N. Pagan-Howard ^19^, L. A. Adler ^20^, S. Alperin ^20^, T. Leon ^20^,Tim B. Bigdeli ^20,EC^, Peter B. Barr^20^, Zoe E. Neale ^20^, Sundar Natarajan ^20^, K. M. Mattocks ^21^, N. Araeva ^21^, J. C. Sullivan ^21^, T. Suppes ^22^, K. Bratcher ^22^, L. Drag ^22^, E.G. Fischer ^22^, L. Fujitani ^22^, S. Gill ^22^, D. Grimm ^22^, J. Hoblyn ^22^, T. Nguyen ^22^, E. Nikolaev ^22^, L. Shere ^22^, R. Relova ^22^, A. Vicencio ^22^, M. Yip ^22^, I. Hurford ^23^, S. Acheampong ^23^, G. Carfagno ^23^, G.L. Haas ^24^, C. Appelt ^24^, E. Brown ^24^, B. Chakraborty ^24^, E. Kelly ^24^, G. Klima ^24^, S. Steinhauer ^24^, R. A. Hurley ^25^, R. Belle ^25^, D. Eknoyan ^25^, K. Johnson ^25^, J. Lamotte ^25^, E. Granholm ^26^, K. Bradshaw ^26^, J. Holden ^26^, R. H. Jones ^26^, T. Le ^26^, I. G. Molina ^26^, M. Peyton ^26^, I. Ruiz ^26^, L. Sally ^26^, J. B. Lohr ^PC,26^, T. Patterson ^PC,26^, A. Tapp ^27^, S. Devroy ^27^, V. Jain ^27^, N. Kilzieh ^27^, L. Maus ^27^, K. Miller ^27^, H. Pope ^27^, A. Wood ^27^, E. Meyer ^28^, P. Givens ^28^, P. B. Hicks ^28^, S. Justice ^28^, K. McNair ^28^, J. L. Pena ^28^, D. F. Tharp ^28^, L. Davis ^29^, M. Ban ^29^, L. Cheatum ^29^, P. Darr ^29^, W. Grayson ^29^, J. Munford ^29^, B. Whitfield ^29^, E. Wilson ^29^, S. E. Melnikoff ^7^, B. L. Schwartz ^7^, M. A. Tureson ^7^, D. D’Souza ^1^, K. Forselius ^1^, John Ko^1^, John Russo^1^, M. Ranganathan ^1^, L. Rispoli ^1^, Yuli Li^1^, Frederick Sayward^1^, Nallakkandi Rajeevan^1^, Krishnan Radhakrishnan^1^, Kei-Hoi Cheung^1^, M. Sather ^8^, C. Colling ^8^, C. Haakenson ^8^, D. Krueger ^8^, Sumitra Muralidhar ^3^, J. Kelsoe ^PC,26^, W. Farwell ^EC,2^, Anil Malhotra ^EC,30^, Polly Palacios ^EC,31^, Thomas Lehner ^PC,32^, Steven R. Marder ^PC,33^, R. Freedman ^PC,34^, Stephen Strakowski ^PC,35^

^EC^ Executive Committee; ^PC^ Planning Committee; ^1^ Clinical Epidemiology Research Center (CERC), VA Connecticut Healthcare System, West Haven, CT; ^2^ Massachusetts Area Veterans Epidemiology, Research, and Information Center (MAVERIC), Jamaica Plain, MA ^3^ Office of Research and Development, Veterans Health Administration, Washington, DC; ^4^ Bruce W. Carter Miami Veterans Affairs (VA) Medical Center, Miami, FL; ^5^ Mental Illness Research, Education and Clinical Center VISN2, James J. Peters VA Medical Center, Bronx, NY; ^6^ Michael E. DeBakey VA Medical Center, Houston, TX ^7^ Washington DC VA Medical Center, Washington, DC; ^8^ Raymond G. Murphy VA Medical Center, Albuquerque, NM; ^9^ Atlanta VA Health Care System, Decatur, GA; ^10^ VA Maryland Health Care System, Baltimore, MD; ^11^ Birmingham VA Medical Center, Birmingham, AL; ^12^ VA Boston Healthcare System, Brockton, MA; ^13^ Ralph H. Johnson VA Medical Center, Charleston, SC; ^14^ Cincinnati VA Medical Center, Cincinnati, OH; ^15^ Richard L. Roudebush VA Medical Center, Indianapolis, IN; ^16^ John L. McClellan Memorial Veterans Hospital, Little Rock, AR; ^17^ VA Long Beach Healthcare System, Long Beach, CA; ^18^ Minneapolis VA Health Care System, Minneapolis, MN; ^19^ VA Hudson Valley Health Care System, Montrose, NY; ^20^ VA New York Harbor Healthcare System, Brooklyn, NY; ^21^ VA Central Western Massachusetts Healthcare System, Leeds, MA; ^22^ VA Palo Alto Health Care System, Palo Alto, CA; ^23^ Corporal Michael J. Crescenz VA Medical Center, Philadelphia, PA; ^24^ VA Pittsburgh Healthcare System, Pittsburgh, PA; ^25^ W.G. (Bill) Hefner VA Medical Center, Salisbury, NC; ^26^ VA San Diego Healthcare System, San Diego, CA; ^27^ VA Puget Sound Health Care System, Tacoma, WA; ^28^ Central Texas Veterans Health Care System, Temple, TX; ^29^ Tuscaloosa VA Medical Center, Tuscaloosa, AL. ^30^ Department of Psychiatry, Zucker Hillside Hospital, Donald and Barbara Zucker School of Medicine at Hofstra/Northwell, Glen Oaks, NY; ^31^ VISN 8, VA Sunshine Healthcare Network, James A. Haley Veterans’ Hospital, Tampa, FL; ^32^ National Institute of Mental Health, Bethesda, MD; ^33^ VA Greater Los Angeles Healthcare System, Los Angeles, CA; ^34^ Department of Psychiatry, University of Colorado School of Medicine, Anschutz Medical Campus,Aurora, CO; ^35^ Department of Psychiatry, University of Texas Dell Medical School, Austin, TX

### Consortium on the Genetics of Schizophrenia (COGS) Investigators

David L Braff, MD ^1,2^, Monica E Calkins, PhD ^3^, Robert R Freedman, MD ^4^, Michael F Green, PhD ^5,6,7^, Tiffany A Greenwood, PhD ^1^, Raquel E Gur, MD, PhD ^3,8-9^, Ruben C Gur, PhD ^3,8-9^, Laura C Lazzeroni, PhD ^10,11^, Gregory A Light, PhD ^1,2^, Keith H Nuechterlein, PhD ^6,7^, Allen D Radant, MD ^12,13^, Larry J Seidman, PhD ^14,15^, Larry J Siever, MD ^16,17^, Jeremy M Silverman, PhD ^16,17^, William S Stone, PhD ^14,15^, Catherine A Sugar, PhD ^18^, Neal R Swerdlow, MD, PhD ^1^, Debby W Tsuang, MD ^12,13^, Ming T Tsuang, MD, PhD, DSc ^1,19,20^, Bruce I Turetsky, MD ^3^

^1^ Department of Psychiatry, University of California, La Jolla, San Diego, CA, USA; ^2^ VISN-22 Mental Illness, Research, Education and Clinical Center (MIRECC), VA San Diego Healthcare System, San Diego, CA, USA; ^3^ Department of Psychiatry, University of Pennsylvania Perelman School of Medicine, Philadelphia, PA, USA; ^4^ Department of Psychiatry, University of Colorado Denver, Aurora, CO, USA; ^5^ VA Greater Los Angeles Healthcare System, Los Angeles, CA, USA; ^6^ Department of Psychiatry and Biobehavioral Sciences and ^7^ Semel Institute for Neuroscience and Human Behavior, Geffen School of Medicine, University of California Los Angeles, Los Angeles, CA, USA; ^8^ Department of Child & Adolescent Psychiatry, University of Pennsylvania Perelman School of Medicine; ^9^ Lifespan Brain Institute, Children’s Hospital of Philadelphia, Philadelphia, PA, USA; ^10^ Departments of Psychiatry and Behavioral Sciences and ^11^ Pediatrics, Stanford University, Stanford, CA, USA; ^12^ Department of Psychiatry and Behavioral Sciences, University of Washington, Seattle, WA, USA; ^13^ VA Puget Sound Health Care System, Seattle, WA, USA; ^14^ Department of Psychiatry, Harvard Medical School, Boston, MA, USA; ^15^ Massachusetts Mental Health Center Public Psychiatry Division of the Beth Israel Deaconess Medical Center, Boston, MA, USA; ^16^ Department of Psychiatry, Icahn School of Medicine at Mount Sinai, NY, USA; ^17^ James J. Peters Veterans Affairs Medical Center, Bronx, NY, USA; ^18^ Department of Biostatistics, University of California Los Angeles School of Public Health, Los Angeles, CA, USA; ^19^ Institute for Genomic Medicine, University of California San Diego, La Jolla, CA, USA; ^20^ Harvard Institute of Psychiatric Epidemiology and Genetics, Boston, MA, USA.

### Genomic Psychiatry Cohort (GPC) Investigators

Michele T Pato MD ^1,2^, Carlos N Pato, MD, PhD ^1,2^, Tim B Bigdeli, PhD ^1,2,3^, Ayman H Fanous, MD ^1,2,3^, Steven A McCarroll, PhD ^4,5^, Peter F Buckley, MD ^6^, Mark J. Daly ^7,8,9,5^, James A Knowles MD, PhD ^2,10^, Douglas S Lehrer, MD ^11^, Dolores Malaspina, MD, MSPH ^12,13^, Mark H Rapaport, MD ^14^, Jeffrey J Rakofsky, MD ^14^, Janet L Sobell, PhD ^15^, Giulio Genovese, PhD ^4,5^, Penelope Georgakopoulos, DrPH ^2^, Jacquelyn L Meyers, PhD ^1^, Roseann E Peterson, PhD ^6^, Helena Medeiros, MSW ^2^, Jorge Valderrama, PhD ^1,2^, Eric D Achtyes, MD ^16^, Roman Kotov, PhD ^17^, Colony Abbott, MPH ^16^, Maria Helena Azevedo, PhD ^18^, Richard A Belliveau, Jr, BA ^4^, Elizabeth Bevilacqua, BS ^19^, Evelyn J Bromet, PhD ^17^, William Byerley, MD ^20^, Celia Barreto Carvalho, PhD ^21^, Sinéad B Chapman, MS ^4^, Lynn E DeLisi, MD ^22,23^, Ashley L Dumont, BASc ^4^, Colm O’Dushlaine, PhD ^4^, Oleg V Evgrafov, PhD ^2,10^, Laura J Fochtmann, MD ^17^, Diane Gage ^4^, James L Kennedy, MD ^24^, Becky Kinkead, PhD ^14^, Antonio Macedo, PhD ^18^, Jennifer L Moran, PhD ^4^, Christopher P Morley, PhD ^25-27^, Mantosh J Dewan, MD ^27^, James Nemesh ^4^, Diana O Perkins, MD, MPH ^28^, Shaun M Purcell, PhD ^4,29^, Edward M Scolnick, MD ^4^, Brooke M Sklar, MA ^15^, Pamela Sklar, MD, PhD ^12,13^, Jordan W Smoller, MD, ScD ^4,23,30,31^, Patrick F Sullivan, MD, FRANZCP ^28,32^, Humberto Nicolini, MD ^33^, Conrad O Iyegbe, PhD ^34^, Fabio Macciardi, MD, PhD ^35^, Stephen R Marder, MD ^36,37^, Michael A Escamilla, MD ^38^, Ruben C Gur, PhD ^39-41^, Raquel E Gur, MD, PhD ^39-41^, Tiffany A Greenwood, PhD ^42^, David L Braff, MD ^42,43^, Marquis P Vawter, PhD, MA, MS ^35^

^1^ Department of Psychiatry and Behavioral Sciences and ^2^ Institute for Genomic Health, SUNY Downstate Medical Center, Brooklyn, NY, USA; ^3^ VA New York Harbor Healthcare System, Brooklyn, NY, USA; ^4^ Stanley Center for Psychiatric Research, Broad Institute of MIT and Harvard, Cambridge, MA, USA; ^5^ Department of Genetics, Harvard Medical School, Boston, MA, USA; ^6^ School of Medicine, Virginia Commonwealth University, Richmond, VA, USA; ^7^ Institute for Molecular Medicine Finland (FIMM), University of Helsinki, Helsinki, Finland; ^8^ Analytic and Translational Genetics Unit, Massachusetts General Hospital, Boston, MA, USA; ^9^ Program in Medical and Population Genetics, Broad Institute of Harvard and MIT, Cambridge, MA, USA; ^10^ Department of Cell Biology, SUNY Downstate Medical Center, Brooklyn, NY, USA; ^11^ Department of Psychiatry, Wright State University, Dayton, OH, USA; ^12^ Departments of Psychiatry and ^13^ Genetics & Genomics, Icahn School of Medicine at Mount Sinai, NY, USA; ^14^ Department of Psychiatry and Behavioral Sciences, Emory University, Atlanta, GA, USA; ^15^ Department of Psychiatry & Behavioral Sciences, University of Southern California, Los Angeles, CA, USA; ^16^ Cherry Health and Michigan State University College of Human Medicine, Grand Rapids, MI, USA; ^17^ Department of Psychiatry, Stony Brook University, Stony Brook, NY, USA; ^18^ Institute of Medical Psychology, Faculty of Medicine, University of Coimbra, Coimbra, PT; ^19^ Beacon Health Options, Boston, MA, USA; ^20^ Department of Psychiatry, University of California, San Francisco, CA, USA; ^21^ Faculty of Social and Human Sciences, University of Azores, PT; ^22^ VA Boston Healthcare System, Brockton, MA, USA; ^23^ Department of Psychiatry, Harvard Medical School, Boston, MA, USA; ^24^ Neurogenetics Laboratory, Campbell Family Mental Health Research Institute, Centre for Addiction and Mental Health; Department of Psychiatry, University of Toronto, ON, CA; ^25^ Departments of Public Health and Preventive Medicine, ^26^ Family Medicine, and ^27^ Psychiatry and Behavioral Sciences, State University of New York, Upstate Medical University, Syracuse, NY, USA; ^28^ Department of Psychiatry, University of North Carolina, Chapel Hill, NC, USA; ^29^ Department of Psychiatry, Brigham and Women’s Hospital, Boston, MA, USA; ^30^ Department of Psychiatry, Massachusetts General Hospital, Boston, MA, USA; ^31^ Department of Epidemiology, Harvard T.H. Chan School of Public Health, Boston, MA, USA; ^32^ Medical Epidemiology and Biostatistics, Karolinska Institutet, Solna, SE; ^33^ Carracci Medical Group, Mexico City, MX; ^34^ Department of Psychosis Studies, King’s College London, London, UK; ^35^ Department of Psychiatry and Human Behavior, University of California, Irvine, CA, USA; ^36^ Department of Psychiatry and Biobehavioral Sciences and ^37^ Semel Institute for Neuroscience and Human Behavior, Geffen School of Medicine, University of California Los Angeles, Los Angeles, CA, USA; ^38^ Department of Psychiatry, University of Texas Rio Grande Valley School of Medicine; ^39^ Departments of Psychiatry and ^40^ Child & Adolescent Psychiatry and ^41^ Lifespan Brain Institute, University of Pennsylvania Perelman School of Medicine and Children’s Hospital of Philadelphia, Philadelphia, PA, USA; ^42^ Department of Psychiatry, University of California, La Jolla, San Diego, CA, USA; ^43^ VISN-22 Mental Illness, Research, Education and Clinical Center (MIRECC), VA San Diego Healthcare System, San Diego, CA, USA

### Project Among African-Americans to Explore Risks for Schizophrenia (PAARTNERS)

Rodney CP Go, PhD ^1^, Raquel Gur, MD, PhD ^2^, Ruben Gur, PhD ^2^, Vish Nimgaonkar, MD, PhD ^3^, Bernive Devlin, PhD ^3^, Monica Calkins, PhD ^2^, Christian Kohler, MD ^2^, Roberta May ^1^; Charlie Swanson, Jr., MD ^1^, Laura Montgomery-Barefield, MD ^1^, Tolulope Aduroja, MD ^1^, Ryan Coleman ^1^; Rakesha Garner ^1^; Lee Prichett, RN ^1^; Thomas Kelley, RN ^1^; Marguerite Ryan Dickson, MS ^1^, Joseph P. McEvoy, MD ^4^, Linda Blalock, RN ^4^, Karen Richardson, MS ^5^, Deirdre Evans-Cosby, MD ^6^, George W. Woods, MD ^6^, Kendaly Meadows, RN ^6^, Sandra Cummings MSW. ^6^, Cara Stephens LCSW ^6^, Kent Baker ^6^, Shirley Hendrix ^7^, Cynthia Gilliard ^7^, Wanda Smalls-Smith ^7^, Steven McLeod-Bryant ^7^, Joel Wood, BS ^3^, Mary Miller, L.P.N. ^3^, Frank Fleischer, M.B.A. ^3^, Kristin Beizai, MD ^8^, Marie Tobin, MD ^8^, Alyssa English, MD ^8^, Richard Sanders, BS ^8^, Shelia A. Dempsey, ADN ^8^, Martha Velez, CSP ^8^, Marianne Smith, BS, MA ^8^, Martha Garriott, MS, NCC ^8^, Nancy Fowler ^8^, Derrick W. Allen, MSSW ^8^, Phyllis Meyer, BS, PA ^8^, Lynn Heustess, BS ^8^

^1^ University of Alabama at Birmingham, Birmingham, AL, USA; ^2^ Department of Child & Adolescent Psychiatry, University of Pennsylvania Perelman School of Medicine, Philadelphia, PA, USA; ^3^ Department of Psychiatry, University of Pittsburgh, Pittsburgh, Pennsylvania, USA; ^4^ Duke University, Durham, NC, USA; ^5^ University of Mississippi, University, MS, USA; ^6^ Morehouse School of Medicine, Atlanta, GA, USA; ^7^ Medical University of South Carolina, Charleston, SC, USA; ^8^ University of Tennessee, Knoxville, TN, USA

### PsychAD Consortium

Aram Hong (1, 4, 6, 7); Athan Z. Li (10, 12); Biao Zeng (1, 4, 6, 7); Chenfeng He (9, 12); Chirag Gupta (9, 12); Christian Porras (1, 4, 6, 7); Clara Casey (1, 4, 6, 7); Colleen A. McClung (18); Collin Spencer (1, 4, 6, 7); Daifeng Wang (9, 10, 12); David A. Bennett (19); David Burstein (1, 2, 4, 6, 7, 8); Deepika Mathur (1, 4, 6, 7); Donghoon Lee (1, 4, 6, 7); Fotios Tsetsos (1, 2, 4, 6, 7); Gabriel E. Hoffman (1, 2, 4, 6, 7, 8); Genadi Ryan (13, 17); Georgios Voloudakis (1, 2, 3, 4, 6, 7, 8); Hui Yang (1, 4, 6, 7); Jaroslav Bendl (1, 4, 6, 7); Jerome J. Choi (11, 12); John F. Fullard (1, 4, 6, 7); Kalpana H. Arachchilage (9, 12); Karen Therrien (1, 4, 6, 7); Kiran Girdhar (1, 4, 6, 7); Lars J. Jensen (21); Lisa L. Barnes (19); Logan C. Dumitrescu (22, 23); Lyra Sheu (1, 4, 6, 7); Madeline R. Scott (18); Marcela Alvia (1, 4, 6, 7); Marios Anyfantakis (1, 4, 6, 7); Maxim Signaevsky (6, 7); Mikaela Koutrouli (1, 4, 6, 7, 21); Milos Pjanic (1, 4, 6, 7); Monika Ahirwar (13, 17); Nicolas Y. Masse (1, 4, 6, 7); Noah Cohen Kalafut (10, 12); Panos Roussos (1, 2, 4, 6, 7, 8); Pavan K. Auluck (20); Pavel Katsel (6); Pengfei Dong (1, 4, 6, 7); Pramod B. Chandrashekar (9, 12); Prashant N.M. (1, 4, 6, 7); Rachel Bercovitch (1, 4, 6, 7); Roman Kosoy (1, 4, 6, 7); Sanan Venkatesh (1, 2, 4, 6, 7); Saniya Khullar (9, 12); Sayali A. Alatkar (10, 12); Seon Kinrot (1, 4, 6, 7); Stathis Argyriou (1, 4, 6, 7); Stefano Marenco (20); Steven Finkbeiner (13, 14, 15, 16, 17); Steven P. Kleopoulos (1, 4, 6, 7); Tereza Clarence (1, 4, 6, 7); Timothy J. Hohman (22, 23); Ting Jin (9, 12); Vahram Haroutunian (5, 6, 7, 8); Vivek G. Ramaswamy (13, 17); Xiang Huang (12); Xinyi Wang (1, 4, 6, 7); Zhenyi Wu (1, 4, 6, 7); Zhiping Shao (1, 4, 6, 7)

### PsychAD Consortium Affiliations

1: Center for Disease Neurogenomics, Icahn School of Medicine at Mount Sinai, New York, NY, USA

2: Center for Precision Medicine and Translational Therapeutics, James J. Peters VA Medical Center, Bronx, NY, USA

3: Department of Artificial Intelligence and Human Health, Icahn School of Medicine at Mount Sinai, New York, NY, USA

4: Department of Genetics and Genomic Sciences, Icahn School of Medicine at Mount Sinai, New York, NY, USA

5: Department of Neuroscience, Icahn School of Medicine at Mount Sinai, New York, NY, USA

6: Department of Psychiatry, Icahn School of Medicine at Mount Sinai, New York, NY, USA

7: Friedman Brain Institute, Icahn School of Medicine at Mount Sinai, New York, NY, USA

8: Mental Illness Research, Education and Clinical Center VISN2, James J. Peters VA Medical Center, Bronx, NY, USA

9: Department of Biostatistics and Medical Informatics, University of Wisconsin-Madison, Madison, WI, USA

10: Department of Computer Sciences, University of Wisconsin-Madison, Madison, WI, USA

11: Department of Population Health Sciences, University of Wisconsin-Madison, Madison, WI, USA

12: Waisman Center, University of Wisconsin-Madison, Madison, WI, USA

13: Center for Systems and Therapeutics, Gladstone Institutes, San Francisco, CA, USA

14: Department of Neurology, University of California San Francisco, San Francisco, CA, USA

15: Department of Physiology, University of California San Francisco, San Francisco, CA, USA

16: Neuroscience and Biomedical Sciences Graduate Programs, University of California San Francisco, San Francisco, CA, USA

17: Taube/Koret Center for Neurodegenerative Disease Research, Gladstone Institutes, San Francisco, CA, USA

18: Department of Psychiatry, University of Pittsburgh School of Medicine, Pittsburgh, PA, USA

19: Rush Alzheimer’s Disease Center and Department of Neurological Sciences, Rush University Medical Center, Chicago, IL, USA

20: Human Brain Collection Core, National Institute of Mental Health-Intramural Research Program, Bethesda, MD, USA

21: Novo Nordisk Foundation Center for Protein Research, Faculty of Health and Medical Sciences, University of Copenhagen, Copenhagen, Denmark

22: Vanderbilt Genetics Institute, Vanderbilt University Medical Center, Nashville, TN, USA

23: Vanderbilt Memory & Alzheimer’s Center, Vanderbilt University Medical Center, Nashville, TN, USA

## Notes

### Competing Interest Statement

The authors have declared no competing interest.

## REFERENCES

1. van der Ven, E. et al. Ethnoracial Risk Variation Across the Psychosis Continuum in the US: A Systematic Review and Meta-Analysis. JAMA Psychiatry 81, 447–455 (2024).

2. Nagendra, A. et al. Neighborhood socioeconomic status and racial disparities in schizophrenia: An exploration of domains of functioning. Schizophr. Res. 224, 95–101 (2020).

3. Anglin, D. M. Racism and Social Determinants of Psychosis. Annu. Rev. Clin. Psychol. 19, 277–302 (2023).

4. International Schizophrenia, Consortium et al. Common polygenic variation contributes to risk of schizophrenia and bipolar disorder. Nature 460, 748–752 (2009).

5. Ikeda, M. et al. Genome-wide association study of schizophrenia in a Japanese population. Biol. Psychiatry 69, 472–478 (2011).

6. de Candia, T. R. et al. Additive genetic variation in schizophrenia risk is shared by populations of African and European descent. Am. J. Hum. Genet. 93, 463–470 (2013).

7. Bigdeli, T. B. et al. Genome-Wide Association Studies of Schizophrenia and Bipolar Disorder in a Diverse Cohort of US Veterans. Schizophr. Bull. (2020) doi:10.1093/schbul/sbaa133.

8. Peterson, R. E. et al. Genome-wide Association Studies in Ancestrally Diverse Populations: Opportunities, Methods, Pitfalls, and Recommendations. Cell 179, 589–603 (2019).

9. Martin, A. R. et al. Human Demographic History Impacts Genetic Risk Prediction across Diverse Populations. Am. J. Hum. Genet. 100, 635–649 (2017).

10. Ding, Y. et al. Polygenic scoring accuracy varies across the genetic ancestry continuum. Nature 618, 774–781 (2023).

11. Duncan, L. et al. Analysis of polygenic risk score usage and performance in diverse human populations. Nat. Commun. 10, 3328 (2019).

12. Bigdeli, T. B. et al. Penetrance and Pleiotropy of Polygenic Risk Scores for Schizophrenia, Bipolar Disorder, and Depression Among Adults in the US Veterans Affairs Health Care System. JAMA Psychiatry (2022).

13. Barr, P. B., Bigdeli, T. B. & Meyers, J. L. Prevalence, Comorbidity, and Sociodemographic Correlates of Psychiatric Disorders Reported in the All of Us Research Program. JAMA Psychiatry 79, 622–628 (2022).

14. Gaziano, J. M. et al. Million Veteran Program: A mega-biobank to study genetic influences on health and disease. J. Clin. Epidemiol. 70, 214–223 (2016).

15. Harvey, P. D. et al. The genetics of functional disability in schizophrenia and bipolar illness: Methods and initial results for VA cooperative study #572. Am. J. Med. Genet. B Neuropsychiatr. Genet. 165B, 381–389 (2014).

16. The ‘All of Us’ Research Program. N. Engl. J. Med. 381, 668–676 (2019).

17. Aliyu, M. H. et al. Project among African-Americans to explore risks for schizophrenia (PAARTNERS): recruitment and assessment methods. Schizophr. Res. 87, 32–44 (2006).

18. Calkins, M. E. et al. The Consortium on the Genetics of Endophenotypes in Schizophrenia: model recruitment, assessment, and endophenotyping methods for a multisite collaboration. Schizophr. Bull. 33, 33–48 (2007).

19. Shi, J. et al. Common variants on chromosome 6p22.1 are associated with schizophrenia. Nature 460, 753–757 (2009).

20. Bigdeli, T. B. et al. Contributions of common genetic variants to risk of schizophrenia among individuals of African and Latino ancestry. Mol. Psychiatry (2019) doi:10.1038/s41380-019-0517-y.

21. Pato, M. T. et al. The genomic psychiatry cohort: partners in discovery. Am. J. Med. Genet. B Neuropsychiatr. Genet. 162B, 306–312 (2013).

22. Calkins, M. E. et al. Project among African-Americans to explore risks for schizophrenia (PAARTNERS): evidence for impairment and heritability of neurocognitive functioning in families of schizophrenia patients. Am. J. Psychiatry 167, 459–472 (2010).

23. Lam, M. et al. Comparative genetic architectures of schizophrenia in East Asian and European populations. Nat. Genet. 51, 1670–1678 (2019).

24. Trubetskoy, V. et al. Mapping genomic loci implicates genes and synaptic biology in schizophrenia. Nature 604, 502–508 (2022).

25. Atkinson, E. G. et al. Tractor uses local ancestry to enable the inclusion of admixed individuals in GWAS and to boost power. Nat. Genet. 53, 195–204 (2021).

26. Bulik-Sullivan, B. K. et al. LD Score regression distinguishes confounding from polygenicity in genome-wide association studies. Nat. Genet. 47, 291–295 (2015).

27. Ge, T., Chen, C.-Y., Ni, Y., Feng, Y.-C. A. & Smoller, J. W. Polygenic prediction via Bayesian regression and continuous shrinkage priors. Nat. Commun. 10, 1776 (2019).

28. Kachuri, L. et al. Principles and methods for transferring polygenic risk scores across global populations. Nat. Rev. Genet. (2023) doi:10.1038/s41576-023-00637-2.

29. Berner, D. Allele Frequency Difference AFD–An Intuitive Alternative to FST for Quantifying Genetic Population Differentiation. Genes 10, 308 (2019).

30. Zhu, K. et al. Multi-omic profiling of the developing human cerebral cortex at the single-cell level. Sci Adv 9, eadg3754 (2023).

31. Corces, M. R. et al. Single-cell epigenomic analyses implicate candidate causal variants at inherited risk loci for Alzheimer’s and Parkinson’s diseases. Nat. Genet. 52, 1158–1168 (2020).

32. Lee, D. et al. Single-cell atlas of transcriptomic vulnerability across multiple neurodegenerative and neuropsychiatric diseases. In Preparation.

33. Zhang, M. J. et al. Polygenic enrichment distinguishes disease associations of individual cells in single-cell RNA-seq data. Nat. Genet. 54, 1572–1580 (2022).

34. Cano-Gamez, E. & Trynka, G. From GWAS to Function: Using Functional Genomics to Identify the Mechanisms Underlying Complex Diseases. Front. Genet. 11, 424 (2020).

35. Mostafavi, H., Spence, J. P., Naqvi, S. & Pritchard, J. K. Systematic differences in discovery of genetic effects on gene expression and complex traits. Nat. Genet. 55, 1866–1875 (2023).

36. Fulco, C. P. et al. Activity-by-contact model of enhancer-promoter regulation from thousands of CRISPR perturbations. Nat. Genet. 51, 1664–1669 (2019).

37. Clarence, T. et al. Simultaneous profiling of transcription and chromatin accessibility at the single-cell level unveils regulatory units critical for postnatal human brain development. In Review.

38. Nasser, J. et al. Genome-wide enhancer maps link risk variants to disease genes. Nature 593, 238–243 (2021).

39. Cochet-Bissuel, M., Lory, P. & Monteil, A. The sodium leak channel, NALCN, in health and disease. Front. Cell. Neurosci. 8, 132 (2014).

40. Wang, K.-S., Liu, X.-F. & Aragam, N. A genome-wide meta-analysis identifies novel loci associated with schizophrenia and bipolar disorder. Schizophr. Res. 124, 192–199 (2010).

41. Souza, R. P. et al. Lack of association of NALCN genetic variants with schizophrenia. Psychiatry Res. 185, 450–452 (2011).

42. Ots, H. D., Tracz, J. A., Vinokuroff, K. E. & Musto, A. E. CD40-CD40L in Neurological Disease. Int. J. Mol. Sci. 23, (2022).

43. Roussos, P. et al. Convergent findings for abnormalities of the NF-κB signaling pathway in schizophrenia. Neuropsychopharmacology 38, 533–539 (2013).

44. Volk, D. W. et al. Molecular mechanisms and timing of cortical immune activation in schizophrenia. Am. J. Psychiatry 172, 1112–1121 (2015).

45. Volk, D. W., Moroco, A. E., Roman, K. M., Edelson, J. R. & Lewis, D. A. The Role of the Nuclear Factor-κB Transcriptional Complex in Cortical Immune Activation in Schizophrenia. Biol. Psychiatry 85, 25–34 (2019).

46. Koopmans, F. et al. SynGO: An Evidence-Based, Expert-Curated Knowledge Base for the Synapse. Neuron 103, 217–234.e4 (2019).

47. Zheutlin, A. B. et al. Penetrance and Pleiotropy of Polygenic Risk Scores for Schizophrenia in 106,160 Patients Across Four Health Care Systems. Am. J. Psychiatry 176, 846–855 (2019).

48. Singh, T. et al. Rare coding variants in ten genes confer substantial risk for schizophrenia. Nature 604, 509–516 (2022).

49. Saunders, G. R. B. et al. Genetic diversity fuels gene discovery for tobacco and alcohol use. Nature 612, 720–724 (2022).

50. Lam, M. et al. Pleiotropic Meta-Analysis of Cognition, Education, and Schizophrenia Differentiates Roles of Early Neurodevelopmental and Adult Synaptic Pathways. Am. J. Hum. Genet. 105, 334–350 (2019).

51. Yaron, A., Huang, P.-H., Cheng, H.-J. & Tessier-Lavigne, M. Differential requirement for Plexin-A3 and -A4 in mediating responses of sensory and sympathetic neurons to distinct class 3 Semaphorins. Neuron 45, 513–523 (2005).

52. Suto, F. et al. Interactions between plexin-A2, plexin-A4, and semaphorin 6A control lamina-restricted projection of hippocampal mossy fibers. Neuron 53, 535–547 (2007).

53. Limoni, G., Murthy, S., Jabaudon, D., Dayer, A. & Niquille, M. PlexinA4-Semaphorin3A-mediated crosstalk between main cortical interneuron classes is required for superficial interneuron lamination. Cell Rep. 34, 108644 (2021).

54. Arbeev, K. G. et al. Interactions between genes involved in physiological dysregulation and axon guidance: role in Alzheimer’s disease. Front. Genet. 14, 1236509 (2023).

55. Suda, S. et al. Decreased expression of axon-guidance receptors in the anterior cingulate cortex in autism. Mol. Autism 2, 14 (2011).

56. Thomas, R. & Yang, X. Semaphorins in immune cell function, inflammatory and infectious diseases. Curr Res Immunol 4, 100060 (2023).

57. Zhang, T. et al. Autophagy collaborates with apoptosis pathways to control oligodendrocyte number. Cell Rep. 42, 112943 (2023).

58. Rusch, H. L. et al. Gene expression differences in PTSD are uniquely related to the intrusion symptom cluster: A transcriptome-wide analysis in military service members. Brain Behav. Immun. 80, 904–908 (2019).

59. Vucicevic, L. et al. Autophagy inhibition uncovers the neurotoxic action of the antipsychotic drug olanzapine. Autophagy 10, 2362–2378 (2014).

60. Metzl, J. M. The Protest Psychosis: How Schizophrenia Became a Black Disease. (Beacon Press, 2010).

61. Metzl, J. M. & Roberts, D. E. Structural Competency Meets Structural Racism: Race, Politics, and the Structure of Medical Knowledge. AMA Journal of Ethics 16, 674–690 (2014).

62. Zhao, B. et al. Common genetic variation influencing human white matter microstructure. Science 372, (2021).

63. Ripke, S. et al. Genome-wide association analysis identifies 13 new risk loci for schizophrenia. Nat. Genet. 45, 1150–1159 (2013).

64. Stevenson, A. et al. Neuropsychiatric Genetics of African Populations-Psychosis (NeuroGAP-Psychosis): a case-control study protocol and GWAS in Ethiopia, Kenya, South Africa and Uganda. BMJ Open 9, e025469 (2019).

65. Morgan, C. et al. Epidemiology of Untreated Psychoses in 3 Diverse Settings in the Global South. JAMA Psychiatry (2022) doi:10.1001/jamapsychiatry.2022.3781.

66. Gitik, M., Bingaman, L. A., Rowland, L. M. & Marques, A. H. The NIMH supports more comprehensive and inclusive genomic studies in psychiatry. World Psychiatry 23, 292–293 (2024).

67. Hunter-Zinck, H. et al. Genotyping Array Design and Data Quality Control in the Million Veteran Program. Am. J. Hum. Genet. 106, 535–548 (2020).

68. Genomic data in the All of Us Research Program. Nature 627, 340–346 (2024).

69. Han, B. & Eskin, E. Random-effects model aimed at discovering associations in meta-analysis of genome-wide association studies. Am. J. Hum. Genet. 88, 586–598 (2011).

70. Lee, S. H. et al. Estimating the proportion of variation in susceptibility to schizophrenia captured by common SNPs. Nat. Genet. 44, 247–250 (2012).

71. Yang, J., Lee, S. H., Goddard, M. E. & Visscher, P. M. GCTA: a tool for genome-wide complex trait analysis. Am. J. Hum. Genet. 88, 76–82 (2011).

72. Wang, G., Sarkar, A., Carbonetto, P. & Stephens, M. A simple new approach to variable selection in regression, with application to genetic fine mapping. J. R. Stat. Soc. Series B Stat. Methodol. 82, 1273–1300 (2020).

73. Zou, Y., Carbonetto, P., Wang, G. & Stephens, M. Fine-mapping from summary data with the ‘Sum of Single Effects’ model. PLoS Genet. 18, e1010299 (2022).

74. Gao, B. & Zhou, X. MESuSiE enables scalable and powerful multi-ancestry fine-mapping of causal variants in genome-wide association studies. Nat. Genet. 56, 170–179 (2024).

75. Yuan, K. et al. Fine-mapping across diverse ancestries drives the discovery of putative causal variants underlying human complex traits and diseases. medRxiv (2023) doi:10.1101/2023.01.07.23284293.

76. Maples, B. K., Gravel, S., Kenny, E. E. & Bustamante, C. D. RFMix: a discriminative modeling approach for rapid and robust local-ancestry inference. Am. J. Hum. Genet. 93, 278–288 (2013).

77. Chang, C. C. et al. Second-generation PLINK: rising to the challenge of larger and richer datasets. Gigascience 4, 7 (2015).

78. de Leeuw, C. A., Mooij, J. M., Heskes, T. & Posthuma, D. MAGMA: generalized gene-set analysis of GWAS data. PLoS Comput. Biol. 11, e1004219 (2015).

79. Barbeira, A. N. et al. Exploring the phenotypic consequences of tissue specific gene expression variation inferred from GWAS summary statistics. Nat. Commun. 9, 1825 (2018).

80. Denny, J. C. et al. PheWAS: demonstrating the feasibility of a phenome-wide scan to discover gene–disease associations. Bioinformatics 26, 1205–1210 (2010).

