## Supplementary Note for "Biological Insights from Schizophrenia-associated Loci in Ancestral Populations"

#### Table of Contents

Cohort Descriptions 2
VA cohorts (Stage 1) 2
Cooperative Studies Program (CSP) #572 2
Million Veteran Program (MVP) 2
Civilian cohorts (Stage 2) 3
All of Us (AOU) Research Program 3
Genomic Psychiatry Cohort (GPC) 3
Consortium on the Genetics of Schizophrenia (COGS) 3
Molecular Genetics of Schizophrenia (MGS) 4
Project Among African-Americans to Explore Risks for Schizophrenia (PAARTNERS) 4
Prevalence of Schizophrenia in the United States 5
The US Veteran Population 5
Cross-biobank comparisons of phenome-wide disease prevalence 8
Supplementary Acknowledgements 9
The CSP #572 study team 9
Million Veteran Program: Consortium Acknowledgement for Manuscripts 10
PsychAD Consortium Authors 12
References 13

### Cohort Descriptions

#### VA cohorts (Stage 1)

##### Cooperative Studies Program (CSP) #572

Participants were recruited through their clinicians, posted notices at participating VA hospitals, and through word of mouth. A total of 27 VA sites participated in this study (with a steady-state of 25 sites). All patients met lifetime DSM-IV criteria for schizophrenia or bipolar I disorder. Patients with major neurologic illnesses, or medical problems that could interfere with central nervous system function, were excluded. Information from medical charts, patients' clinicians, or other informants were used, if needed, to confirm diagnoses— with all participants receiving the Structured Clinical Interview for the DSM (SCID). [1] Diagnosed substance abuse was not an exclusion criterion, given the high level of co-occurrence in the population and concerns about representativeness.

##### Million Veteran Program (MVP)

Participants are active users of the VHA healthcare system, and were recruited through invitational mailings or by MVP staff while receiving clinical care. Informed consent and authorization per the Health Insurance Portability and Accountability Act were the only other inclusion criteria. All participants complete a Baseline Survey which includes information on demographic factors, family pedigree, health status, lifestyle habits, military experiences, medical history, family history of specific illnesses and physical features; and may also complete an optional Lifestyle Survey. [2] At the time of writing, 4,697 individuals (~50% of CSP #572; ~0.7% of MVP) were dually enrolled in both studies.

We compiled relevant ICD-9/10 codes for schizophrenia, schizoaffective disorder, and bipolar disorders, and lists of commonly prescribed antipsychotics and mood stabilizers, and extracted all matching records from the VA Corporate Data Warehouse (CDW). We required at least two inpatient or outpatient billing codes for schizophrenia. For individuals who met this criterion for schizophrenia and bipolar illness, we gave preeminence to the "prevailing" diagnosis, i.e. the mode of the diagnoses recorded during the five most recent clinical encounters. We selected control participants by leveraging the EHR, after exclusion of individuals with any history of diagnosis of schizophrenia, schizoaffective disorders, psychosis, or bipolar disorder, past treatment with antipsychotics or mood-stabilizing medications, or who self-endorsed a personal or family history of either condition on the MVP core demographics survey.

CSP #572 and MVP participants were genotyped on the MVP 1.0 Axiom array. [3] Genetic ancestry of participants was classified using the HARE method, [4] which harmonizes genetic ancestry with self-identified race and ethnicity.

### Civilian cohorts (Stage 2)

#### All of Us (AOU) Research Program

The All of Us Research Program is a prospective, nationwide cohort study of the effects of lifestyle, environment, and genomics on health outcomes. Participant recruitment is predominantly done through participating health care organizations and in partnership with Federally Qualified Health Centers. Participants can enroll as direct volunteers, visiting community-based enrollment sites. Enrollment, informed consent, and baseline health surveys are done digitally through the All of Us website (https://joinallofus.org). Participants are then invited to undergo a basic physical examination, biospecimen collection, and follow-up via linkage with EHRs and periodic surveys. We included data from participants enrolled between May 6, 2018, and July 1, 2022 who had available EHR and short-read WGS data, excluding individuals who self-reported ever having held active-duty status in the US military in order to avoid any sample overlaps with MVP (Release 7, N = 187,358). To determine case-control status, we emulated procedures described above for MVP, further integrating SNOMED terms. [5] Applying the prevailing diagnosis definition for schizophrenia, a strict definition for screened controls, and limiting to African genetic ancestry yielded 1,065 cases and 27,866 screened controls. All analyses were conducted in accordance with the All of Us Code of Conduct.

#### Genomic Psychiatry Cohort (GPC)

The Genomic Psychiatry Cohort (GPC) is a cosmopolitan sample of cases and controls living in local communities and being treated in healthcare delivery systems. All participants enrolled as probable cases were interviewed using the Diagnostic Interview for Psychosis and Affective Disorders (DI-PAD), a semi-structured clinical interview developed specifically for the GPC study using the same principles as those applied in the development of the Diagnostic Interview for Psychosis – Diagnostic Module (DIP-DM), [6] and incorporated questions developed for the Diagnostic Interview for Genetic Studies (DIGS). [7] In the present analysis, all cases met lifetime diagnostic criteria for schizophrenia or schizoaffective disorder (depressed type) in accordance with the OPCRIT algorithms for DSM-IV and/or ICD-10 [8] criteria, and/or DSM-5. [9] Individuals reporting no lifetime symptoms indicative of psychosis or mania and who have no first-degree relatives with these symptoms were included as control participants. Exclusion criteria included any premorbid organic mental disorders (i.e., epilepsy, CNS infection, significant head trauma, mental retardation), and premorbid history of significant drug or alcohol dependence by DSM IV/5 that confounds the diagnosis of schizophrenia. Genotyping was performed in four waves using the Illumina Omni2.5M array and Global Screening Array (GSA). [10,11]

#### Consortium on the Genetics of Schizophrenia (COGS)

The Consortium on the Genetics of Schizophrenia (COGS) a multi-site study which recruited outpatients diagnosed with schizophrenia or schizoaffective disorder, depressed type, as well as healthy control participants. Recruitment for COGS was conducted in two phases across multiple academic sites, including the University of California (San Diego and Los Angeles), Icahn School of Medicine at Mount Sinai, University of Pennsylvania, University of Washington, University of Colorado Health Sciences Center, and Harvard University. Recruitment sources included clinician referrals, outpatient facilities, local National Alliance for the Mentally Ill chapters, and media advertisements. Standardized psychiatric interviews using the SCID were conducted for all participants to obtain consensus diagnoses. All schizophrenia patients met DSM-IV criteria; healthy control participants

(HCPs) were excluded for psychosis, bipolar disorder, current depression, substance abuse, or a first-degree relative with a history of psychosis. Participants ranged from 18 to 65 years of age and underwent urine toxicology screens before assessment. HCPs were matched to patients based on age, sex, ancestry, and parental educational attainment. Participants with severe systemic illness, significant head injury, neurological disorders, positive drug or alcohol screens, recent substance abuse or dependence, or an estimated premorbid IQ below 70 were excluded. Written informed consent was obtained from each participant after a detailed description of the study. The study received ethics approval from the local Institutional Review Boards at each site. AFR ancestry participants were genotyped using Illumina Multi-Ethnic Global Array (MEGA) array at the Broad Institute.[12,13]

#### Molecular Genetics of Schizophrenia (MGS)

Participants in the Molecular Genetics of Schizophrenia (MGS) study were recruited from multiple sites in the USA and Australia, as detailed in previous studies. [14] Participants provided written informed consent, and the human subjects protocol was approved by the Institutional Review Boards (IRBs) at each collection site. All cases met DSM-IV criteria for schizophrenia or schizoaffective disorder. Genotyping of the DNA samples was conducted at the Broad Institute using the Affymetrix 6.0 array. Individual-level data were accessed via dbGAP (phs000167.v1.p1; phs000021.v3.p2).

#### Project Among African-Americans to Explore Risks for Schizophrenia (PAARTNERS)

The Project among African-Americans to explore risks for schizophrenia (PAARTNERS) included probands meeting DSM-IV criteria for schizophrenia or schizoaffective disorder depressed type, their relatives (as trios, affected sibling pairs, or multiplex pedigrees), and community controls.[15,16] Participants were recruited in the Southeastern and Eastern USA, self-identified as African American, and were assessed using the Diagnostic Interview for Genetic Studies, Family Interview for Genetic Studies, the Penn-CNB, and medical records review. Blood or buccal cells were collected for genetic analyses. Probands and ancestry-matched, psychiatrically unscreened controls from MiGEN [17] were genotyped on the MEGA.

### Prevalence of Schizophrenia in the United States

In the United States, higher rates of diagnosed schizophrenia in persons racialized as Black and those of African ancestries is a well documented phenomenon, and has been attributed to differences in access to care, cultural or racial bias in diagnosis, and chronic stress arising from structural racism. [18–21] Evidence from several European countries similarly highlights increased rates of psychiatric conditions among immigrant, migrant, and asylum seeking persons, especially men. [22,23] Identifying the causal and contributing factors underlying disparities in incidence and outcome is challenging, requiring robust epidemiological data that is not confounded by ascertainment and selection biases. [24,25]

#### The US Veteran Population

The Veterans Health Administration (VHA) is the largest integrated healthcare system in the United States, offering comprehensive, no-cost, and centrally managed care to 18 million veterans, with around 9 million individuals currently enrolled. These services are available to individuals with prior active-duty military service (eligibility for which begins at 18 years of age) that meet certain criteria. The MVP cohort (*N*=815,516 at the time of writing) is representative of the population that utilizes VHA services: 90.7% are men who at study enrollment averaged 62.6 years of age (standard deviation, SD=13.9); among 75,811 female veterans, the average age is 50.4 years (SD=13.6). Around 46.5% of participants reported having served during the Vietnam War (1964-1975), which coincides with the compulsory enlistment of individuals into the armed forces known as "the draft." Notably, we observe increasing numbers of Black men born during this period, who later enrolled in VHA services following completion of their military service (**Supplementary Figure 1**).

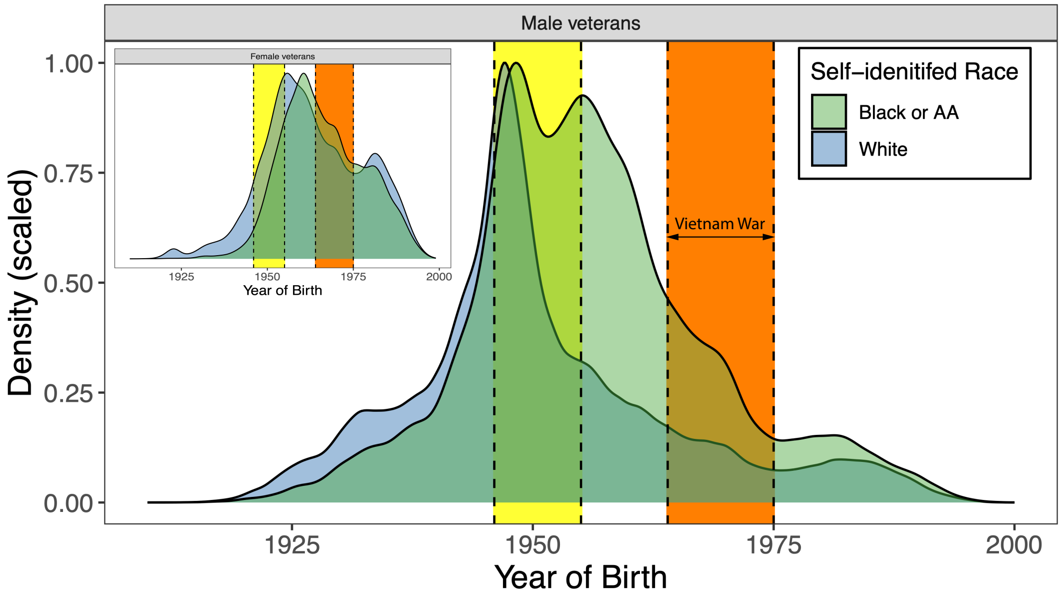


**Supplementary Figure 1. Birth year by self-identified race and gender.** Males born between 1946 and 1955 (displayed by yellow highlighted region) reached their eighteenth birthday during the active period of the Vietnam War (1964-1975; displayed by orange highlighted region), thus becoming newly eligible for the draft. The corresponding distributions for female veterans are displayed in the inset plot.

Given its well-defined age group and being composed primarily of young men who underwent screening for military service, the draft-era population is an advantageous setting for studying the onset and outcomes of illnesses that emerge in early adulthood. The age of onset of schizophrenia in men is typically between 18 and 25 years, with earlier manifestations often characterized by worse premorbid functioning that could preclude entry into military service. [26] Adverse exposures and experiences can precipitate or worsen psychotic symptoms, and are also more common among veterans (both during and following their military service) than in the general population. With these epidemiological considerations in mind, we reexamined disparities in schizophrenia diagnosis here, newly integrating self-report information on participants' prior military service and gender.

The overall lifetime prevalence of schizophrenia (based on 2 or more ICD-9/10 codes) in MVP is around 2.7%; among individuals who self-identified as Black or African American, this rose to 5.6%, compared to 1.9% among self-identified White individuals. However, among Black men who served during the Vietnam war, the prevalence was 6.5%, which further increased to 7.1% in the years following (**Supplementary Figure 2**). Importantly, we also observed a higher prevalence among White male veterans during these periods, and in both groups, rates of diagnosed schizophrenia have declined in the 1990s, reaching 2.6% and 1.3% for Black and White men who served in the last two decades. Intriguingly, we observed a similar trend among Black and White female veterans, who were not subject to the draft, but for whom recognition and representation in the military increased during the Vietnam war and following decades.

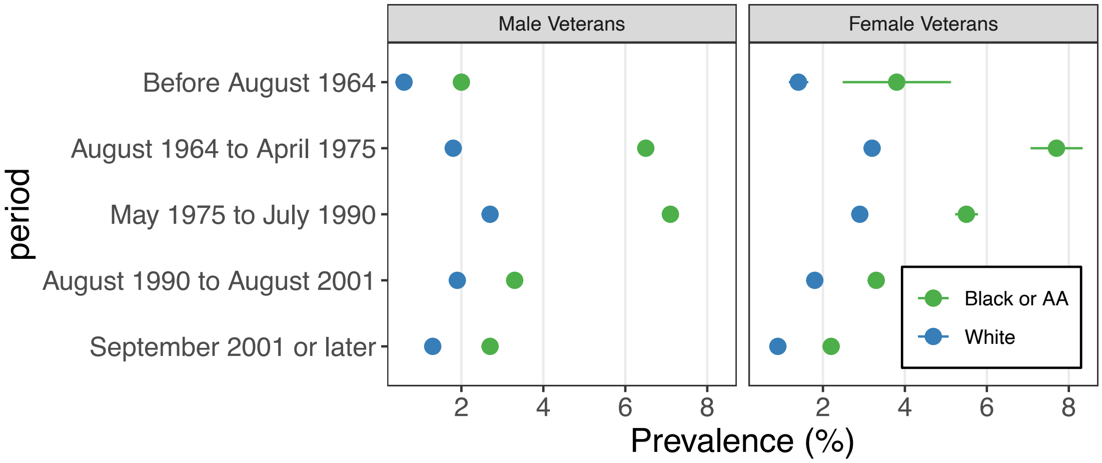


**Supplementary Figure 2. Prevalence by self-reported race, gender, and military service period.** For MVP participants who self-identified as Male, Female, Black, and White, prevalence estimates and standard errors are displayed. In total, *N*=103,439 individuals reported serving after September 2001; *N*=203,647 from 1990 to 2001, *N*=189,321 from 1975 to 1990, *N*=378,854 from 1964 to 1975, and *N*=137,627 before 1964.

Overall, these observations indicate that within the VHA, racial disparities in schizophrenia diagnoses have diminished in recent decades, potentially as a consequence of transitioning to a volunteer force, resulting in selection into military service by healthier individuals. It is also worth noting that our results may be conservative, given our requirement that cases have at least two ICD-9/10 billing codes for schizophrenia; results for inpatient settings and any lifetime diagnosis of schizophrenia (or broader psychosis) might yield different results.

Next, we asked whether the relationship between an individual's genetic liability and their lifetime risk of being diagnosed with schizophrenia might differ across settings, collapsing the service periods above into broad 30-year epochs. Among individuals of AFR genetic ancestry whose service occurred between 1964 and 1990, we observe a 5% prevalence of schizophrenia in the lowest decile of risk, compared to 10% for those in the highest decile (**Supplementary Figure 3**). During the same period, ~1% of EUR participants in the bottom decile and nearly 5% of those in the top decile had received a diagnosis. We observe a near-identical relationship among EUR individuals in later periods. In stark contrast, we observe a drop in absolute prevalence among AFR persons after 1990, with prevalence rates of 2% and 5% in the lowest and highest strata. We compared results for a body mass index (BMI) PRS and obesity (two or more ICD-9/10 codes), observing relatively consistent PRS effects across epochs, but less marked population differences in absolute prevalence.

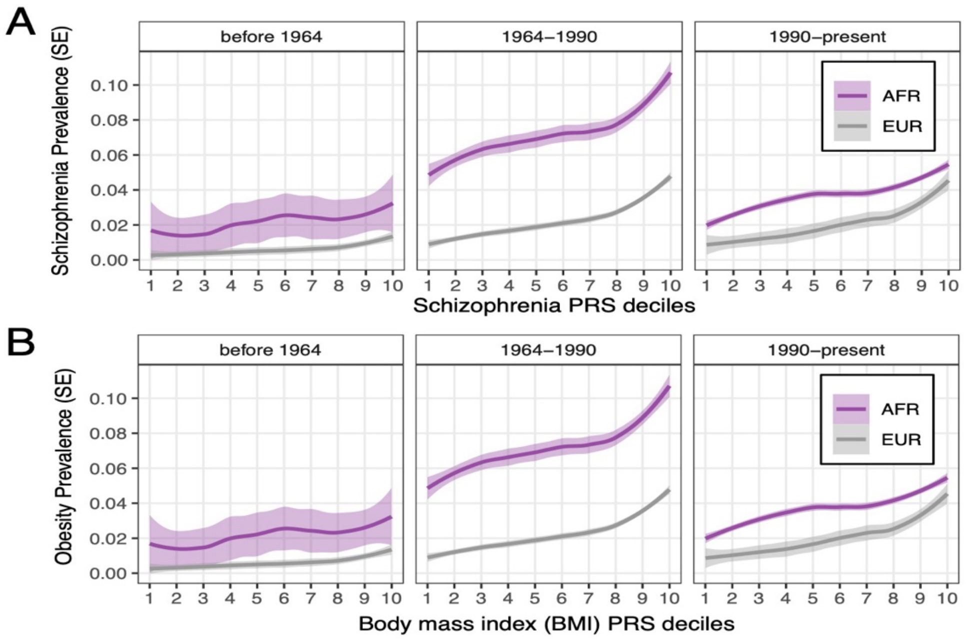


**Supplementary Figure 3. Case prevalence by PRS decile and service period. A)** For each decile of schizophrenia PRS, the prevalence of schizophrenia (PheCode 295.1) is shown **B)** For each decile of BMI PRS, the prevalence of obesity (Phecode 278.1) is shown. individuals were assigned to ancestry-specific PRS deciles, adjusted for 10 principal components (PCs), age, and sex; we then re-estimated the prevalence of schizophrenia across deciles for each service period. Shaded areas give an estimate's standard error.

Our results support a consistent and replicable association between schizophrenia PRS and diagnosis that is largely independent of context and ancestry. However, they cannot account for the relative contributions of PRS, unmeasured biological factors, and individuals' unique environmental risk and resilience factors. Furthermore, these results do not discount factors related to clinician biases, [19] adverse exposures before, during, or following military service, or self-selection for VHA service utilization. To gain a better understanding of these persistent population-based disparities, future research should emphasize collection of additional exposure and experiential data, and detailed clinical evaluation of symptoms over the course of early illness in representative samples.

#### Cross-biobank comparisons of phenome-wide disease prevalence

We have previously reported that the proportion of individuals that met criteria for schizophrenia in the AOU Research Program cohort is around 0.9%, using a definition of 2+ phecodes. [5] Here, we specifically excluded participants who reported having been active-duty military, and observed that among the remaining AFR individuals, the lifetime proportion of schizophrenia diagnoses was 4.1%. We extended our comparisons of prevalence phenome-wide, estimating the correlation between log-scale estimates for each biobank (**Supplementary Figure 4**). Using 170 ontologically distinct Phecodes with prevalence of 1% in both cohorts, we calculated a Pearson's correlation of 0.63 ($P$=2.86×10$^{-20}$) between AOU and MVP. We performed the same comparison for MVP-AFR and MVP-EUR, using 288 phecodes with prevalence of 1% or greater in both ancestries, and obtained an estimate of 0.94 ($P$=1.49×10$^{-132}$).

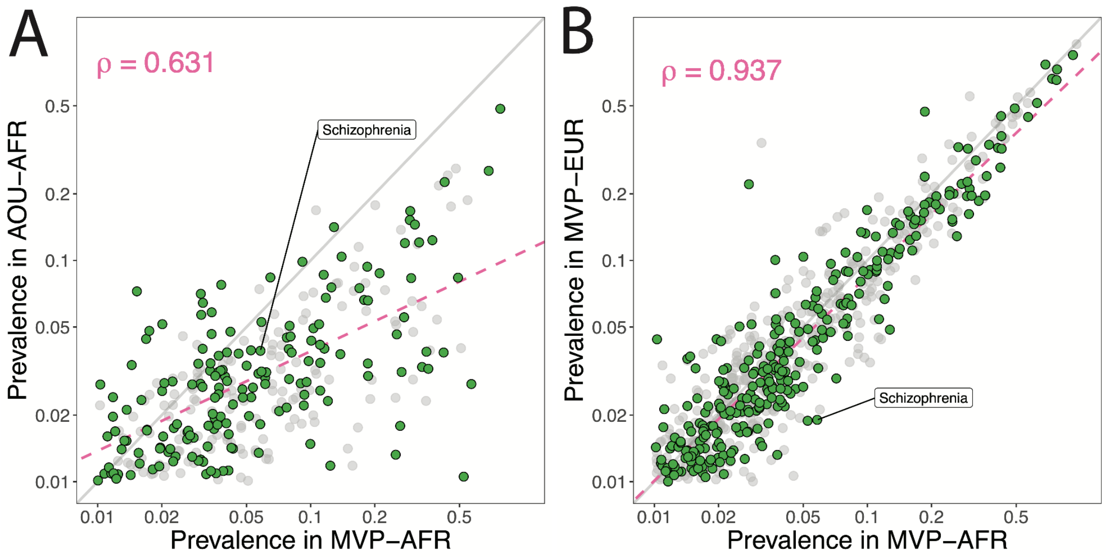


**Supplementary Figure 4. Phenome-wide prevalence differences for MVP and AOU. A)** For overlapping PheWAS terms in MVP-AFR and AOU-AFR results, the prevalence in each population is displayed on the $\log_{10}$ scale. **B)** For overlapping terms in MVP-AFR and MVP-EUR PheWAS results, scaled prevalence estimates are displayed. Initial correlations were estimated from the **green** highlighted points. **Pink** dashed lines indicate the regression of prevalence estimates onto MVP-AFR data.The identity line is shown in **gray**.

We repeated this procedure 10,000 times for each comparison, retaining one phecode per parent term, and from the resultant set, randomly sampling 100 codes from which to re-estimate the correlation; this yielded average estimates of 0.62 (standard deviation, SD=0.08) and 0.93 (SD=0.02) for comparisons of MVP-AFR with AOU-AOU and MVP-EUR, respectively.

That in spite of the noted disparities in schizophrenia diagnoses, we observed a stronger overall correspondence between prevalence estimates for AFR and EUR participants in MVP compared to AFR participants in MVP and AOU is noteworthy. This supports the advantages of the VHA as a setting for population-scale epidemiological research, while also highlighting the challenges of harmonizing EHRs across healthcare settings,[27] which may compound biases in representation arising from selection, utilization, and population demographics.

### Supplementary Acknowledgements

#### The CSP #572 study team

**Planning Committee:** M. Aslan, M. Antonelli, M. de Asis, M. S. Bauer, M. Brophy, J. Concato, F. Cunningham, R. Freedman, M. Gaziano, T. Gleason, P. D. Harvey, G. Huang, J. Kelsoe, T. Kosten, T. Lehner, J. B. Lohr, S. R. Marder, P. Miller, T.J. O'Leary, T. Patterson, P. Peduzzi, R. Przygodzki, L. Siever, P. Sklar, S. Strakowski, H. Zhao.
**Executive Committee:** M. Brophy, J. Concato, A.H. Fanous, W. Farwell, M. Gaziano, P.D. Harvey, T. Kosten, A. Malhotra, S. Mane, P. Palacios, P. Sklar, L. Siever, H. Zhao.
**Study Chairs' Offices:** VA Healthcare System, Bronx, NY, included L. Siever (Study Co- Chair), M. Corsey, L. Zaluda. VA Healthcare System, Miami, FL, included P.D. Harvey (Study Co- Chair), J. Johnson.
**CSP Epidemiology Centers:** VA Clinical Epidemiology Research Center (CERC), VA Connecticut Healthcare System, West Haven, CT, included J. Concato (Director, Methodological Co-Principal Proponent), M. Aslan, D. Cavaliere, V. Jeanpaul, A. Maffucci, L. Mancini, Y. Li, N. Rajeevan, J. Ko, J. Russo, F. Sayward, K.H. Cheung, K. Radhakrishnan; the Massachusetts Veterans Epidemiology Research and Information Center (MAVERIC), VA Boston Healthcare System, Jamaica Plain, MA, included M. Gaziano (Director, Methodological Co-Principal Proponent), J. Deen, G. Muldoon, S. Whitbourne.
**Study Sites:** *Albuquerque*: J. Canive, L. Adamson, L. Calais, G. Fuldauer, R. Kushner, G. Toney, M. Lackey, A. Mank, N. Mahdavi, G. Villarreal. *Atlanta*: E. C. Muly, F. Amin, M. Dent, J. Wold. *Baltimore*: B. Fischer, A. Elliott, C. Felix, G. Gill. *Birmingham*: P. E. Parker, C. Logan, J. McAlpine. *Brockton*: L.E. DeLisi, S. G Reece. *Charleston*: M.B. Hammer, D. Agbor-Tabie, W. Goodson. *Cincinnati*: M. Aslam, M. Grainger, Neil Richtand, Alexander Rybalsky. *Houston*: R. Al Jurdi, E. Boeckman, T. Natividad, D. Smith, M. Stewart, S. Torres, Z. Zhao. *Indianapolis*: A. Mayeda, A. Green, J. Hofstetter, S. Ngombu, M. K. Scott, A. Strasburger, J. Sumner. *Little Rock*: G. Paschall, J. Mucciarelli, R. Owen, S. Theus, D. Tompkins. *Long Beach*: S.G. Potkin, C. Reist, M. Novin, S. Khalaghizadeh. *Miami*: R. Douyon, J. Johnson, N. Kumar, B. Martinez. *Minneapolis*: S.R. Sponheim, T.L. Bender, H.L. Lucas, A.M. Lyon, M.P. Marggraf, L.H. Sorensen, C.R. Surerus. *Montrose*: C. Sison, J. Amato, D.R. Johnson, N. Pagan-Howard. *New York Harbor*: L.A. Adler, S. Alperin, T. Leon. *Northampton*: K.M. Mattocks, N. Araeva, J.C. Sullivan. *Palo Alto*: T. Suppes, K. Bratcher, L. Drag, E.G. Fischer, L. Fujitani, S. Gill, D. Grimm, J. Hoblyn, T. Nguyen, E. Nikolaev, L. Shere, R. Relova, A. Vicencio, M. Yip. *Philadelphia*: I. Hurford, S. Acheampong, G. Carfagno. *Pittsburgh*: G.L. Haas, C. Appelt, E. Brown, B. Chakraborty, E. Kelly, G. Klima, S. Steinhauer. *Salisbury*: R.A. Hurley, R. Belle, D. Eknoyan, K. Johnson, J. Lamotte. *San Diego*: E. Granholm, K. Bradshaw, J. Holden, R. H. Jones, T. Le, I.G. Molina, M. Peyton, I. Ruiz, L. Sally. *Tacoma*: A. Tapp, S. Devroy, V. Jain, N. Kilzieh, L. Maus, K. Miller, H. Pope, A. Wood. *Temple*: E. Meyer, P. Givens, P. B. Hicks, S. Justice, K. McNair, J.L. Pena, D.F. Tharp. *Tuscaloosa*: L. Davis, M. Ban, L. Cheatum, P. Darr, W. Grayson, J. Munford, D. Smith, B. Whitfield, E. Wilson. *Washington DC*: A.H. Fanous, S.E. Melnikoff, B.L Schwartz, M.A. Tureson. *West Haven*: D. D'Souza, K. Forselius, M. Ranganathan, L. Rispoli.
**Albuquerque, NM, CSP Coordinating Center (for monitoring)**: M. Sather (Director), C. Colling, C. Haakenson, D. Krueger.
**VA Office of Research and Development**: T. O'Leary (Chief Research and Development Officer), G. Huang (Director, Cooperative Studies Program), T. Gleason (Director, Clinical Science Research and Development Service), R. Przygodzki (Associate Director for Genomic Medicine, and Acting Director of Biomedical Laboratory Research and Development Service), S. Muralidhar (Senior Scientific Program Manager Genomic Medicine Program, Biomedical and Clinical R&D Services).

### Million Veteran Program: Consortium Acknowledgement for Manuscripts

**MVP Executive Committee**
- Co-Chair: J. Michael Gaziano, M.D., M.P.H.
- Co-Chair: Rachel Ramoni, D.M.D., Sc.D.
- Jim Breeling, M.D. ex-officio
- Kyong-Mi Chang, M.D.
- Grant Huang, Ph.D.
- Sumitra Muralidhar, Ph.D.
- Christopher J. O'Donnell, M.D., M.P.H.
- Philip S. Tsao, Ph.D.

**MVP Program Office**
- Sumitra Muralidhar, Ph.D.
- Jennifer Moser, Ph.D.

**MVP Recruitment/Enrollment**
- Recruitment/Enrollment Director/Deputy Director, Boston
- Stacey B. Whitbourne, Ph.D.; Jessica V. Brewer, M.P.H.
- MVP Coordinating Centers

o Clinical Epidemiology Research Center CERC, West Haven – John Concato, M.D., M.P.H.
o Cooperative Studies Program Clinical Research Pharmacy Coordinating Center, Albuquerque - Stuart Warren, J.D., Pharm D.; Dean P. Argyres, M.S.
o Genomics Coordinating Center, Palo Alto – Philip S. Tsao, Ph.D.
o Massachusetts Veterans Epidemiology Research Information Center MAVERIC, Boston - J. Michael Gaziano, M.D., M.P.H.
o MVP Information Center, Canandaigua – Brady Stephens, M.S.

- Core Biorepository, Boston – Mary T. Brophy M.D., M.P.H.; Donald E. Humphries, Ph.D.
- MVP Informatics, Boston – Nhan Do, M.D.; Shahpoor Shayan
- Data Operations/Analytics, Boston – Xuan-Mai T. Nguyen, Ph.D.

**MVP Science**
- Genomics - Christopher J. O'Donnell, M.D., M.P.H.; Saiju Pyarajan Ph.D.; Philip S. Tsao, Ph.D.
- Phenomics - Kelly Cho, M.P.H, Ph.D.
- Data and Computational Sciences – Saiju Pyarajan, Ph.D.
- Statistical Genetics – Elizabeth Hauser, Ph.D.; Yan Sun, Ph.D.; Hongyu Zhao, Ph.D.

**MVP Local Site Investigators**
- Atlanta VA Medical Center Peter Wilson - Bay Pines VA Healthcare System Rachel McArdle
- Birmingham VA Medical Center Louis Dellitalia
- Cincinnati VA Medical Center John Harley
- Clement J. Zablocki VA Medical Center Jeffrey Whittle
- Durham VA Medical Center Jean Beckham
- Edith Nourse Rogers Memorial Veterans Hospital John Wells
- Edward Hines, Jr. VA Medical Center Salvador Gutierrez
- Fayetteville VA Medical Center Gretchen Gibson
- VA Health Care Upstate New York Laurence Kaminsky
- New Mexico VA Health Care System Gerardo Villareal
- VA Boston Healthcare System Scott Kinlay
- VA Western New York Healthcare System Junzhe Xu

- Ralph H. Johnson VA Medical Center Mark Hamner
- Wm. Jennings Bryan Dorn VA Medical Center Kathlyn Sue Haddock
- VA North Texas Health Care System Sujata Bhushan
- Hampton VA Medical Center Pran Iruvanti
- Hunter Holmes McGuire VA Medical Center Michael Godschalk
- Iowa City VA Health Care System Zuhair Ballas
- Jack C. Montgomery VA Medical Center Malcolm Buford
- James A. Haley Veterans' Hospital Stephen Mastorides
- Louisville VA Medical Center Jon Klein
- Manchester VA Medical Center Nora Ratcliffe
- Miami VA Health Care System Hermes Florez
- Michael E. DeBakey VA Medical Center Alan Swann
- Minneapolis VA Health Care System Maureen Murdoch
- N. FL/S. GA Veterans Health System Peruvemba Sriram
- Northport VA Medical Center Shing Shing Yeh
- Overton Brooks VA Medical Center Ronald Washburn
- Philadelphia VA Medical Center Darshana Jhala
- Phoenix VA Health Care System Samuel Aguayo
- Portland VA Medical Center David Cohen
- Providence VA Medical Center Satish Sharma
- Richard Roudebush VA Medical Center John Callaghan
- Salem VA Medical Center Kris Ann Oursler
- San Francisco VA Health Care System Mary Whooley
- South Texas Veterans Health Care System Sunil Ahuja
- Southeast Louisiana Veterans Health Care System Amparo Gutierrez
- Southern Arizona VA Health Care System Ronald Schifman
- Sioux Falls VA Health Care System Jennifer Greco
- St. Louis VA Health Care System Michael Rauchman
- Syracuse VA Medical Center Richard Servatius
- VA Eastern Kansas Health Care System Mary Oehlert
- VA Greater Los Angeles Health Care System Agnes Wallbom
- VA Loma Linda Healthcare System Ronald Fernando
- VA Long Beach Healthcare System Timothy Morgan
- VA Maine Healthcare System Todd Stapley
- VA New York Harbor Healthcare System Scott Sherman
- VA Pacific Islands Health Care System Gwenevere Anderson
- VA Palo Alto Health Care System Philip Tsao
- VA Pittsburgh Health Care System Elif Sonel
- VA Puget Sound Health Care System Edward Boyko
- VA Salt Lake City Health Care System Laurence Meyer
- VA San Diego Healthcare System Samir Gupta
- VA Southern Nevada Healthcare System Joseph Fayad
- VA Tennessee Valley Healthcare System Adriana Hung
- Washington DC VA Medical Center Jack Lichy
- W.G. Bill Hefner VA Medical Center Robin Hurley
- White River Junction VA Medical Center Brooks Robey
- William S. Middleton Memorial Veterans Hospital Robert Striker

### PsychAD Consortium Authors

Aram Hong (1, 4, 6, 7); Athan Z. Li (10, 12); Biao Zeng (1, 4, 6, 7); Chenfeng He (9, 12); Chirag Gupta (9, 12); Christian Porras (1, 4, 6, 7); Clara Casey (1, 4, 6, 7); Colleen A. McClung (18); Collin Spencer (1, 4, 6, 7); Daifeng Wang (9, 10, 12); David A. Bennett (19); David Burstein (1, 2, 4, 6, 7, 8); Deepika Mathur (1, 4, 6, 7); Donghoon Lee (1, 4, 6, 7); Fotios Tsetsos (1, 2, 4, 6, 7); Gabriel E. Hoffman (1, 2, 4, 6, 7, 8); Genadi Ryan (13, 17); Georgios Voloudakis (1, 2, 3, 4, 6, 7, 8); Hui Yang (1, 4, 6, 7); Jaroslav Bendl (1, 4, 6, 7); Jerome J. Choi (11, 12); John F. Fullard (1, 4, 6, 7); Kalpana H. Arachchilage (9, 12); Karen Therrien (1, 4, 6, 7); Kiran Girdhar (1, 4, 6, 7); Lars J. Jensen (21); Lisa L. Barnes (19); Logan C. Dumitrescu (22, 23); Lyra Sheu (1, 4, 6, 7); Madeline R. Scott (18); Marcela Alvia (1, 4, 6, 7); Marios Anyfantakis (1, 4, 6, 7); Maxim Signaevsky (6, 7); Mikaela Koutrouli (1, 4, 6, 7, 21); Milos Pjanic (1, 4, 6, 7); Monika Ahirwar (13, 17); Nicolas Y. Masse (1, 4, 6, 7); Noah Cohen Kalafut (10, 12); Panos Roussos (1, 2, 4, 6, 7, 8); Pavan K. Auluck (20); Pavel Katsel (6); Pengfei Dong (1, 4, 6, 7); Pramod B. Chandrashekar (9, 12); Prashant N.M. (1, 4, 6, 7); Rachel Bercovitch (1, 4, 6, 7); Roman Kosoy (1, 4, 6, 7); Sanan Venkatesh (1, 2, 4, 6, 7); Saniya Khullar (9, 12); Sayali A. Alatkar (10, 12); Seon Kinrot (1, 4, 6, 7); Stathis Argyriou (1, 4, 6, 7); Stefano Marenco (20); Steven Finkbeiner (13, 14, 15, 16, 17); Steven P. Kleopoulos (1, 4, 6, 7); Tereza Clarence (1, 4, 6, 7); Timothy J. Hohman (22, 23); Ting Jin (9, 12); Vahram Haroutunian (5, 6, 7, 8); Vivek G. Ramaswamy (13, 17); Xiang Huang (12); Xinyi Wang (1, 4, 6, 7); Zhenyi Wu (1, 4, 6, 7); Zhiping Shao (1, 4, 6, 7)

**PsychAD Consortium Affiliations**

1: Center for Disease Neurogenomics, Icahn School of Medicine at Mount Sinai, New York, NY, USA
2: Center for Precision Medicine and Translational Therapeutics, James J. Peters VA Medical Center, Bronx, NY, USA
3: Department of Artificial Intelligence and Human Health, Icahn School of Medicine at Mount Sinai, New York, NY, USA
4: Department of Genetics and Genomic Sciences, Icahn School of Medicine at Mount Sinai, New York, NY, USA
5: Department of Neuroscience, Icahn School of Medicine at Mount Sinai, New York, NY, USA
6: Department of Psychiatry, Icahn School of Medicine at Mount Sinai, New York, NY, USA
7: Friedman Brain Institute, Icahn School of Medicine at Mount Sinai, New York, NY, USA
8: Mental Illness Research, Education and Clinical Center VISN2, James J. Peters VA Medical Center, Bronx, NY, USA
9: Department of Biostatistics and Medical Informatics, University of Wisconsin-Madison, Madison, WI, USA
10: Department of Computer Sciences, University of Wisconsin-Madison, Madison, WI, USA
11: Department of Population Health Sciences, University of Wisconsin-Madison, Madison, WI, USA
12: Waisman Center, University of Wisconsin-Madison, Madison, WI, USA
13: Center for Systems and Therapeutics, Gladstone Institutes, San Francisco, CA, USA
14: Department of Neurology, University of California San Francisco, San Francisco, CA, USA
15: Department of Physiology, University of California San Francisco, San Francisco, CA, USA
16: Neuroscience and Biomedical Sciences Graduate Programs, University of California San Francisco, San Francisco, CA, USA
17: Taube/Koret Center for Neurodegenerative Disease Research, Gladstone Institutes, San Francisco, CA, USA
18: Department of Psychiatry, University of Pittsburgh School of Medicine, Pittsburgh, PA, USA
19: Rush Alzheimer's Disease Center and Department of Neurological Sciences, Rush University Medical Center, Chicago, IL, USA
20: Human Brain Collection Core, National Institute of Mental Health-Intramural Research Program, Bethesda, MD, USA
21: Novo Nordisk Foundation Center for Protein Research, Faculty of Health and Medical Sciences, University of Copenhagen, Copenhagen, Denmark
22: Vanderbilt Genetics Institute, Vanderbilt University Medical Center, Nashville, TN, USA
23: Vanderbilt Memory & Alzheimer's Center, Vanderbilt University Medical Center, Nashville, TN, USA

### References


1. First, M. B., Spitzer, R. L., Gibbon, M. & Williams, J. B. W. *User's Guide for the Structured Clinical Interview for DSM-IV Axis I Disorders SCID-I: Clinician Version*. (American Psychiatric Pub, 1997).
2. Gaziano, J. M. *et al.* Million Veteran Program: A mega-biobank to study genetic influences on health and disease. *J. Clin. Epidemiol.* **70**, 214–223 (2016).
3. Hunter-Zinck, H. *et al.* Genotyping Array Design and Data Quality Control in the Million Veteran Program. *Am. J. Hum. Genet.* **106**, 535–548 (2020).
4. Fang, H. *et al.* Harmonizing Genetic Ancestry and Self-identified Race/Ethnicity in Genome-wide Association Studies. *Am. J. Hum. Genet.* **105**, 763–772 (2019).
5. Barr, P. B., Bigdeli, T. B. & Meyers, J. L. Prevalence, Comorbidity, and Sociodemographic Correlates of Psychiatric Disorders Reported in the All of Us Research Program. *JAMA Psychiatry* **79**, 622–628 (2022).
6. Castle, D. J. *et al.* The diagnostic interview for psychoses (DIP): development, reliability and applications. *Psychol. Med.* **36**, 69–80 (2006).
7. Nurnberger, J. I. *et al.* Diagnostic Interview for Genetic Studies: Rationale, Unique Features, and Training. *Arch. Gen. Psychiatry* **51**, 849–859 (1994).
8. World Health Organization. *The ICD-10 Classification of Mental and Behavioural Disorders: Diagnostic Criteria for Research*. (World Health Organization, 1993).
9. American Psychiatric Association. *Diagnostic and Statistical Manual of Mental Disorders (DSM-5®)*. (American Psychiatric Pub, 2013).
10. Pato, M. T. *et al.* The genomic psychiatry cohort: partners in discovery. *Am. J. Med. Genet. B Neuropsychiatr. Genet.* **162B**, 306–312 (2013).
11. Bigdeli, T. B. *et al.* Contributions of common genetic variants to risk of schizophrenia among individuals of African and Latino ancestry. *Mol. Psychiatry* (2019) doi:10.1038/s41380-019-0517-y.
12. Calkins, M. E. *et al.* The Consortium on the Genetics of Endophenotypes in Schizophrenia: model recruitment, assessment, and endophenotyping methods for a multisite collaboration. *Schizophr. Bull.* **33**, 33–48 (2007).
13. Light, G. *et al.* Comparison of the heritability of schizophrenia and endophenotypes in the COGS-1 family study. *Schizophr. Bull.* **40**, 1404–1411 (2014).
14. Shi, J. *et al.* Common variants on chromosome 6p22.1 are associated with schizophrenia. *Nature* **460**, 753–757 (2009).
15. Aliyu, M. H. *et al.* Project among African-Americans to explore risks for schizophrenia (PAARTNERS): recruitment and assessment methods. *Schizophr. Res.* **87**, 32–44 (2006).
16. Calkins, M. E. *et al.* Project among African-Americans to explore risks for schizophrenia (PAARTNERS): evidence for impairment and heritability of neurocognitive functioning in families of schizophrenia patients. *Am. J. Psychiatry* **167**, 459–472 (2010).

17. O’Donnell, C. J. *et al.* Genome-wide association study for coronary artery calcification with follow-up in myocardial infarction. *Circulation* **124**, 2855–2864 (2011).
18. Heun-Johnson, H. *et al.* Association Between Race/Ethnicity and Disparities in Health Care Use Before First-Episode Psychosis Among Privately Insured Young Patients. *JAMA Psychiatry* **78**, 311–319 (2021).
19. Gara, M. A., Minsky, S., Silverstein, S. M., Miskimen, T. & Strakowski, S. M. A Naturalistic Study of Racial Disparities in Diagnoses at an Outpatient Behavioral Health Clinic. *Psychiatr. Serv.* **70**, 130–134 (2019).
20. Metzl, J. M. & Roberts, D. E. Structural Competency Meets Structural Racism: Race, Politics, and the Structure of Medical Knowledge. *AMA Journal of Ethics* **16**, 674–690 (2014).
21. Metzl, J. M. *The Protest Psychosis: How Schizophrenia Became a Black Disease*. (Beacon Press, 2010).
22. Termorshuizen, F. *et al.* The incidence of psychotic disorders among migrants and minority ethnic groups in Europe: findings from the multinational EU-GEI study. *Psychol. Med.* **52**, 1376–1385 (2022).
23. Tarricone, I. *et al.* Migration history and risk of psychosis: results from the multinational EU-GEI study. *Psychol. Med.* **52**, 2972–2984 (2022).
24. Anderson, K. K., McKenzie, K. J. & Kurdyak, P. Examining the impact of migrant status on ethnic differences in mental health service use preceding a first diagnosis of schizophrenia. *Soc. Psychiatry Psychiatr. Epidemiol.* **52**, 949–961 (2017).
25. Gureje, O. & Cohen, A. Differential outcome of schizophrenia: where we are and where we would like to be. *Br. J. Psychiatry* **199**, 173–175 (2011).
26. Harvey, P. D. *et al.* Clinical, cognitive and functional characteristics of long-stay patients with schizophrenia: a comparison of VA and state hospital patients. *Schizophr. Res.* **43**, 3–9 (2000).
27. Zheutlin, A. B. *et al.* Penetrance and Pleiotropy of Polygenic Risk Scores for Schizophrenia in 106,160 Patients Across Four Health Care Systems. *Am. J. Psychiatry* **176**, 846–855 (2019).
